# Psilocybin, cognitive stability and drinking reduction in co-occurring alcohol use disorder and depression: a pilot secondary analysis of a randomized, placebo-controlled trial

**DOI:** 10.64898/2026.10.27.26366630

**Authors:** Ceyda Sayalı, Eli Weisman, Frederick S. Barrett

**Author notes:** **Corresponding author:** Ceyda Sayalı, PhD, Center for Psychedelic and Consciousness Research, Johns Hopkins University School of Medicine.

## Abstract

**Background:** Change in cognitive control is a candidate transdiagnostic mechanism of psychedelic therapy. One account predicts that, once acute effects resolve, psilocybin shifts control toward flexibility while sparing inhibition and working memory. A motivational account (PRISM) predicts that psilocybin increases the willingness to exert control rather than the ability to do so. We tested both in patients with co-occurring alcohol use disorder (AUD) and major depression.

**Methods:** In this secondary analysis of a randomized, double-blind, placebo-controlled trial of psilocybin for AUD with comorbid major depressive disorder (MDD; ClinicalTrials.gov NCT04620759), a subset of participants (placebo n=13, psilocybin n=12) completed five cognitive tasks before drug administration and again about one week after it (mean 6.7 days, SD 1.1). Outcome measures spanned the stability–flexibility continuum. Free choice between levels of cognitive demand indexed the participant’s willingness to exert control. Effect sizes and Bayes factors were used to estimate drug effects and associations with change in drinking and depressive symptoms across 34 retest measures.

**Results:** Relative to placebo, psilocybin slowed responding in cued task switching (learning-phase RT d=1.01, BF10=7.7; switch trials d=1.12, BF10=17). Psilocybin effects on feedback-driven measures were inconsistent in direction (7 of 11 toward flexibility); the largest was on perseveration, which rose less with practice under psilocybin than under placebo (d=−0.79, BF10=1.1). Other drug effects on the 34 retest measures had inconclusive Bayes factors (BF10=0.40–2.4), and none survived false discovery rate correction. Demand avoidance, analyzed separately from the 34 measures, showed no detectable drug effect (BF01=2.0). Baseline demand avoidance showed no detectable relation to performance but tracked depressive symptoms (*r_s_*=.71, BF10=24). Drinking fell in both arms, and on 15 of 17 stability–flexibility measures change toward stability accompanied drinking reduction (stability composite *r_s_*=−.64, BF10=29; *r^s^*=−.43 when change was adjusted for baseline scores). Lower baseline working-memory span predicted larger drinking reductions (BF10=19–25). Depressive symptoms fell more after psilocybin than after placebo (PHQ-9, d=−0.88, BF10=2.7), but their change was not associated with cognitive change or predicted by working-memory span.

**Conclusions:** About one week after dosing, psilocybin slowed cued task switching but had no consistent effect on feedback-driven flexibility, and there was no evidence of an effect on the willingness to exert effort. Drinking reduction was associated with increased, not reduced, cognitive stability, whereas improvement in depressive symptoms was not associated with cognitive change. These pilot estimates can guide adequately powered trials.

## Introduction

Alcohol use disorder (AUD) and major depression (MDD) frequently co-occur, and each worsens the course of the other (Boden & Fergusson, 2011). Psilocybin-assisted therapy has shown promise for both: it reduced heavy drinking relative to placebo in AUD (Bogenschutz et al., 2022) and produced rapid and sustained antidepressant effects in patients with MDD and treatment-resistant depression (Goodwin et al., 2022; Gukasyan et al., 2022). However, effects of psilocybin on reducing drinking in patients with AUD are mixed (Rieser et al., 2025). Mechanisms by which psilocybin may reduce either depression or alcohol use remain unclear. Without a mechanistic account of these effects, it is difficult to predict who will benefit, or whether a single mechanism can underlie therapeutic response in disorders as different as AUD and depression.

Cognitive control is a candidate shared therapeutic target of psychedelics, and change in control a candidate mechanism of their effects, because deficits in control processes are observed in patients with both depression and substance use disorders (Goschke, 2014; McTeague et al., 2016). Control is not a single function, however, but a family of related processes, and psychedelics need not change all of them in the same way. The unity/diversity model distinguishes response inhibition, working-memory updating and set shifting, which share a common factor but dissociate behaviorally and neurally (Miyake et al., 2000; Miyake & Friedman, 2012). Control can also be described by the balance it strikes between stability and flexibility (Goschke & Bolte, 2014; Sayalı & Barrett, 2023). Stability maintains a goal and shields it from interference; flexibility updates goals and strategies when circumstances change. Each has a cost at its extreme: too much stability produces rigid, perseverative behavior, and too much flexibility produces distractibility and goal neglect.

One account of the cognitive mechanisms underlying psychedelic effects proposed that psychedelics act on control in a component-specific and time-dependent way (Sayalı et al., 2026). During the acute psychedelic experience, inhibition, working memory and structured set shifting are impaired. Once acute effects resolve, flexibility is selectively enhanced while inhibition and working memory return to baseline. The clearest clinical evidence for this account comes from a study of patients with MDD (Doss et al., 2021). This study found fewer perseverative errors on a card-sorting task one week and one month after two doses of psilocybin, with no observed change in inhibition performance during a Stroop task. This has been interpreted as a recalibration of the stability–flexibility balance toward a more exploratory mode, with reduced perseveration and greater behavioral adaptability after intervention (Sayalı et al., 2026). This leads to two open questions: does the same hold for other forms of flexibility, such as task switching, which have not been systematically studied? And, does the early post-acute period, in the first days after dosing, differ from later time periods?

Performance on a control task reflects how much control a person deploys. That depends both on what they are able to do and on how much control they choose to invest, which is set by weighing the expected benefits of control against its subjective costs (Shenhav et al., 2013; Botvinick & Braver, 2015). The willingness to invest control (motivation) can be measured separately from the capacity to exert that control by comparing choices between tasks that differ in demand (Westbrook et al., 2013; Sayalı & Badre, 2019), and motivation dissociates from performance behaviorally and neurally (Westbrook et al., 2020; Sayalı & Badre, 2019, 2021). In patients with depression, reduced willingness to invest effort can occur even when the ability to perform is preserved (Grahek et al., 2019). Building on this distinction, the PRISM model (Psychedelic Recalibration of Internal Salience and Mental Effort Costs; Sayalı, 2026) proposes that post-acute cognitive change after psychedelics reflects a recalibrated valuation of effort, with lower perceived costs and greater salience of meaningful goals, rather than increased capacity. Sayalı (2026) names the Demand Selection Task, in which participants choose freely between levels of demand, as a direct test. The PRISM model predicts that psilocybin will increase the willingness to choose demanding tasks without a corresponding change in capacity, and that people with high effort avoidance but preserved performance will benefit most from psychedelic intervention. It also states a boundary condition: the increase is expected mainly for personally meaningful goals.

AUD adds a further complication, because the direction of change that would support recovery is not obvious. Some accounts of substance use disorder emphasize excessive stability, in the form of habitual responding to alcohol cues that resists change (Everitt & Robbins, 2016; Sjoerds et al., 2013). Others emphasize insufficient stability, in the form of impulsive choice and weak maintenance of abstinence goals (MacKillop et al., 2011). When AUD co-occurs with depression, it is unclear which direction along the continuum supports reduced drinking. Drinking also often falls substantially in the placebo arms of trials in patients with AUD (Litten et al., 2013). Therefore, recovery may not depend on drug-induced cognitive change at all. Rather, individual differences in cognitive function may predict who benefits from treatment regardless of arm (Bates et al., 2013).

We examined these questions with a five-task battery administered before and about one week after double-blind drug administration in a randomized, placebo-controlled trial of psilocybin for co-occurring AUD and MDD. The battery sampled components of the unity/diversity model of cognitive control. Inhibition was measured with the Multi-Source Interference Task (MSIT). Shifting was measured in two forms: probabilistic rule switching, in which rule changes must be inferred from feedback, and cued task switching, in which the task is signaled on every trial. Value-based learning and the stability of choice were estimated with a computational model of probabilistic reward and punishment learning, and working memory with operation span. The cued task-switching paradigm was the Parametric Demand Selection Task (Sayalı & Badre, 2019), which also offers free choice between demand levels and therefore provides the battery’s only measure of the willingness to exert control. Rather than selecting a few summary scores, we placed every measure on the stability–flexibility continuum according to its construct definition (Table 1) and examined the battery as a whole. Because every task was repeated, practice effects are expected on every measure; the placebo arm provides the reference against which drug effects are judged.

**Table 1.** Position of each measure on the stability–flexibility continuum, with score type. Performance scores reflect control as deployed, which depends on both ability and the level of control that a participant invests; only free choice between demand levels indexes the willingness to engage control directly. The 34 retest measures comprise 30 performance and 4 choice measures; demand avoidance was analyzed separately.

| Task | Measure | Score type | Stability–flexibility: a higher score means |
| --- | --- | --- | --- |
| PRL | Reward-learning accuracy; punishment-learning accuracy | Performance | Neither (value learning) |
| PRL | Learning rate (gain, loss) | Performance (model-derived) | Higher flexibility: faster updating of values |
| PRL | Inverse temperature (gain, loss) | Performance (model-derived) | Stability: exploitation of learned values |
| PRL | Perseveration | Performance (model-derived) | Stability: repeating the previous choice regardless of outcome |
| Rule switching | Overall accuracy; median RT (learning, transfer) | Performance | Neither |
| Rule switching | Switch cost (learning, transfer) | Performance | Stability: slower adaptation after a rule change |
| Rule switching | Pre-switch accuracy (learning, transfer) | Performance | Stability: maintenance of the current rule |
| Rule switching | Post-switch accuracy (learning, transfer) | Performance | Flexibility: adaptation to a new rule |
| Demand selection | Accuracy and RT (learning, decision phase) | Performance | Neither |
| Demand selection | Task-switch cost in accuracy and RT (learning, decision phase) | Performance | Stability: task-set inertia |
| Demand selection | Demand avoidance (primary); P(least demanding), P(most demanding), mean chosen level; selection RT | Motivation (choice) | Neither |
| MSIT | Control and interference accuracy and RT | Performance | Neither |
| MSIT | Interference effect (RT, accuracy) | Performance | Flexibility: weaker goal shielding |
| Operation span | Partial-credit span (baseline) | Performance | Neither |

We asked four questions. First, does psilocybin move feedback-driven and cued shifting toward flexibility relative to placebo, while leaving inhibition unchanged? Second, does psilocybin increase the willingness to exert control, and is that willingness separable from performance? Third, does change in stability, flexibility or willingness to exert control track change in drinking and depressive symptoms, and in which direction? Fourth, do baseline working-memory span or demand avoidance predict who reduces drinking, and does this depend on the drug?

## Methods

### Participants and design

This is a secondary analysis of a randomized, double-blind, placebo-controlled trial of psilocybin in adults with AUD and comorbid MDD (ClinicalTrials.gov NCT04620759). Twenty-five participants completed baseline cognitive testing (placebo n=13, psilocybin n=12). Twenty-three also completed post-dose cognitive testing (two placebo participants completed only baseline testing and contributed only to baseline analyses). Baseline testing took place a median of 1 day before double-blind drug administration (range 1–8 days), and post-dose testing a mean of 6.7 days after it (SD 1.1; median 7, range 4–9; median 7 days in each arm). The two testing sessions were a median of 8 days apart (range 5–13). Sample characteristics are given in Table 2.

**Table 2.** Baseline characteristics of the participants with cognitive data.

| Characteristic | Placebo (n=13) | Psilocybin (n=12) |
| --- | --- | --- |
| Age, years, mean (SD) | 41.8 (10.1) | 42.3 (9.8) |
| Female, n | 5 | 8 |
| Taking an antidepressant, n | 4 | 6 |
| QIDS-SR at screening, mean (SD) | 17.5 (3.2) | 15.9 (2.4) |
| PHQ at baseline visit, mean (SD) [n] | 7.5 (3.4) [11] | 9.2 (3.7) [12] |
| Proportion of drinking days, baseline 90 days | .66 (.24) | .57 (.24) |
| Proportion of heavy-drinking days | .41 (.25) | .38 (.22) |
| Drinks per drinking day | 6.1 (3.0) | 6.1 (2.5) |
| Operation span, mean (SD) [n] | 10.0 (2.6) [10] | 9.8 (2.3) [8] |

### Cognitive battery

Tasks were administered in a fixed order within each session: emotional memory practice, the Parametric Demand Selection Task (learning phase, then decision phase), emotional memory encoding, probabilistic reward learning, MSIT, operation span, probabilistic rule switching and emotional memory retrieval. The emotional memory task is not reported here.

#### Probabilistic reward learning (PRL)

Participants were presented with a series of trials containing two figures (cues) on a computer screen. Participants were asked to choose between two abstract cues on gain trials (win or no win) and loss trials (loss or no loss). In each pair, one cue led to the better outcome on 75% of trials (Pessiglione et al., 2006). A training run was followed by two test runs with new cue pairs (112 responded test trials; missed trials were re-presented and excluded). Accuracy was calculated as the proportion of choices of the better cue, separately for gain and loss trials. Choices were fitted to a Rescorla–Wagner model with a softmax choice rule, separate learning rates and inverse temperatures for gain and loss trials, and a perseveration parameter that biases choice toward repeating the previous choice regardless of its outcome. Values were reset at the start of each run and trial type, and parameters were estimated by maximum likelihood (best of 50 random starts). Adding perseveration improved AIC (mean ΔAIC=−2.5, t(46)=−2.70, p=.010, over all 47 sessions) but not BIC (p=.78).

#### Probabilistic rule switching

This task was adapted from the low-volatility condition of Wen et al. (2023; https://github.com/tanya-wen/Meta-flexibility). Participants categorized images by one of two stimulus dimensions, guided by feedback that was valid on 80% of trials. The relevant dimension changed without warning. A 120-trial learning phase (rule change every 30 trials) was followed by a 120-trial transfer phase with two new dimensions (rule change every 20 trials). Accuracy was computed for each phase on the five trials before each rule change (pre-switch accuracy) and the five trials after it (post-switch accuracy), averaged over the three (learning) or five (transfer) changes. Switch cost was calculated as pre-switch minus post-switch accuracy.

#### Parametric Demand Selection Task

This task followed Sayalı and Badre (2019; github.com/zceydas/ParametricEffortSelectionTask). Participants were presented with a sequence of individual playing cards (one per trial). Their task was to make cued parity (e.g. is the number even or odd?) or magnitude (e.g. is the number greater or less than 5) judgments, with the task signaled on every trial. Demand was manipulated as the probability of a task switch (.13, .38, .63 or .88), each associated with a card deck (e.g. the color of the back of a given deck) that was displayed at the beginning of each block of trials (each block containing 8 trials). In a learning phase (320 trials), participants performed every demand level. In a decision phase (72 trials), they chose freely between two decks on each trial and then performed the chosen deck. No external incentive was attached to this choice. Performance was indexed by accuracy, response time (RT), and task-switch costs (switch minus repeat trials) in each phase. The willingness to exert control was indexed by demand avoidance, defined as the proportion of choices of the less demanding deck across all pairings (0.5 = indifference; Sayalı & Badre, 2019). The probabilities of choosing the least and the most demanding deck, the mean chosen level, and selection RT are reported as secondary choice measures.

#### MSIT

On each trial, three digits were shown, and participants identified the digit that differed from the other two (Bush & Shin, 2006). On control trials, the other two digits were zeros and the target appeared in the position matching its value; on interference trials, the other two digits were non-zero and the target’s position did not match its value. Eight alternating blocks of control and interference trials yielded 192 trials. Interference effects (interference minus control) were computed for RT (correct trials) and accuracy.

#### Operation span

The automated operation span task (Unsworth et al., 2005) was scored from trial-level data as the sum, across 15 test sets, of correctly recalled letters divided by set size (range 0–15). Primary analyses used baseline scores from all valid sessions; post-dose scores, available for a subset, are reported in the Supplement (Table S8).

### Classifying measures on the stability–flexibility continuum

Each measure was classified by its position on the stability–flexibility continuum (Table 1). Scores also differ in what they can reveal about control. Every score other than free choice is a performance score: it reflects control as deployed, which depends jointly on ability and on how much control the participant chooses to invest (Shenhav et al., 2013). Performance scores therefore cannot isolate capacity. Free choice between demand levels without incentives indexes the willingness to engage control more directly (Sayalı & Badre, 2019), although choices can still be shaped by how demanding each level feels. We use "motivation" only for that choice measure. Because demand avoidance is the battery’s only measure of motivation, score type is shown in Table 1 but not used as a classification dimension; demand avoidance is analyzed separately, as the test of PRISM’s prediction. Following convention, we refer to operation span, a performance score, as an index of working-memory capacity, with the same caveat.

On the continuum, stability measures index the maintenance of a current choice, rule or task set: perseveration, high inverse temperature (exploitation of learned values), switch costs, and pre-switch accuracy. Flexibility measures index updating and openness to change: higher learning rates, post-switch accuracy, and MSIT interference effects. Interference effects index susceptibility to task-irrelevant information and can be read as weaker shielding of the goal; flexibility in the sense of adapting to relevant change is captured by learning rates and post-switch accuracy. Overall accuracy, speed, span, and choice measures were not assigned a pole. For model parameters, the pole refers to the direction of the scale: lower learning rates and higher inverse temperatures indicate greater stability. Placements followed construct definitions rather than results, but the classification was finalized after data collection, so all analyses that depend on it are exploratory.

### Data quality

Missed PRL trials were excluded before model fitting. In the demand selection task, missed responses and presses of non-task keys were counted as errors and excluded from RT. In the same task, the baseline sessions of six participants whose learning-phase accuracy was not above chance (binomial p≥.05 over 320 trials; accuracy .48–.54) were excluded from all measures of that task, because deck–demand associations could not have been learned. In the rule-switching task, three baseline sessions and one post-dose session whose overall accuracy did not exceed chance (binomial p≥.05 over 240 trials; accuracy .45–.53) were excluded from all measures of that task, because rule-dependent indices cannot be interpreted when the rule was not followed; results with these sessions included, and with the stricter criterion of Wen et al. (2023; overall accuracy below 65%), are reported as sensitivity analyses. In two post-dose sessions, decision-phase responses were inverted on 95–96% of answered trials, consistent with a reversed key mapping, and accuracy was scored accordingly. In the MSIT, the key mapping was determined per block as the rule that reproduced at least 70% of responses (one block with a mid-session change of mapping could not be resolved and was not scored); two post-dose sessions in which 98–99% of interference trials were answered by the target’s position rather than its identity were excluded. Details are given in the Supplementary Methods.

### Clinical measures

Drinking was assessed with the Timeline Followback (Sobell & Sobell, 1992) at baseline (90 days leading up to screening) and over the 30 and 90 days after experimental drug administration. Because the windows differed in length, drinking was expressed as the proportion of drinking days, the proportion of heavy-drinking days, and the number of drinks per drinking day. Heavy-drinking days were days with five or more drinks for men and four or more for women. The primary drinking outcome was the change from baseline to the 30-day window, which contained the post-dose cognitive session. Drinking over time was compared between arms with a linear mixed model of each drinking proportion with arm, window (baseline, 30 days and 90 days after experimental drug administration) and their interaction as fixed effects with a random intercept per participant. Depressive symptoms were measured with the 9-item Patient Health Questionnaire (PHQ-9; Kroenke et al., 2001) at the baseline and post-dose cognitive visits, and with the 16-item Quick Inventory of Depressive Symptomatology–Self-Report (QIDS-SR; Rush et al., 2003) at screening.

### Statistical analysis

#### Drug effects

Analysis of covariance (ANCOVA) was used to estimate the effect of allocation (psilocybin vs placebo) on post-dosing score when controlling for baseline score for each of the 34 retest measures (Vickers & Altman, 2001). Effects are reported as the baseline-adjusted arm difference in pooled-SD units (d; psilocybin minus placebo) with 95% confidence intervals, and p values were corrected for the false discovery rate (FDR) across the 34 tests (Benjamini & Hochberg, 1995); FDR-adjusted p values are reported as q. Within-arm change was tested with paired t-tests. As a sensitivity analysis, the number of days between drug administration and post-dose testing was added as a covariate.

#### Position on the continuum

For the 17 measures with an assigned pole, drug effects and clinical couplings were re-signed so that positive values indicate movement toward stability. Consistency of direction was summarized with sign tests, which are descriptive because measures from the same task are correlated. As a summary index, we computed a stability composite from the principal stability measure of each feedback-driven task: perseveration and transfer-phase switch cost. Change in each measure (post-dose minus baseline) was z-scored across all participants, with both arms pooled, and the composite was the mean of the available z-scores, so that higher values indicate movement toward stability. Participants with only one of the two measures contributed that measure. Because rule-switching change was available for fewer participants, 13 of the 22 participants contributed both measures and 9 contributed perseveration only; analyses of the composite were therefore repeated in the 13 participants with both measures. The composite was compared between arms with Welch’s t-test. As a sensitivity analysis, a broader composite was computed by averaging eight indices spanning all four tasks (perseveration, mean inverse temperature, mean learning rate, both rule-switching costs, both cued RT switch costs and the MSIT RT interference effect), each z-scored as previously described, aligned so that positive values indicate movement toward stability (flexibility-pole indices sign-reversed), and requiring at least three included measures per participant. Because the arms differed at baseline on transfer-phase switch cost, the stability composite was also recomputed from baseline-adjusted change, with each measure’s post-dose score residualized on its baseline score (pooled across arms) before z-scoring.

#### Baseline construct validation

To test whether the classification captured individual differences that are shared across measures, we correlated scores across measures assessed at the baseline session, which preceded randomization and is independent of drug effects or clinical outcomes. We selected one index per construct and task: perseveration, mean inverse temperature and mean learning rate (PRL); pre-switch and post-switch accuracy averaged over phases (rule switching, sessions above chance); task-switch costs in RT and in accuracy averaged over phases (demand selection, sessions above chance); and the MSIT RT interference effect. Each index was aligned so that higher values indicate greater stability, with flexibility-pole indices sign-reversed. If the classification is valid, aligned indices from different tasks should correlate positively. We tested the mean Spearman correlation across the 23 cross-task pairs against a null distribution from 10,000 permutations in which participants were shuffled independently within each index, preserving missing data. Within-task pairs were excluded from the test because they share trials or, for model parameters, estimation trade-offs. Test–retest reliability of each index was estimated as the Spearman correlation between baseline and post-dose scores after removing arm means. The classification was not revised in light of these results. Factor analysis was not attempted, because only 12 participants contributed all eight indices.

#### Associating change in cognition with clinical outcomes

For every cognitive measure, change (post minus pre) was related to change in the proportion of drinking days with Spearman correlations (*r_s_*; all correlations are Spearman unless stated otherwise) pooled across arms, with rank-based regression adjusting for arm, and with a rank-based regression that added an arm × change interaction. FDR correction was applied across the 34 couplings. The same models, and the same re-signing and sign test on the continuum, were applied to change in PHQ-9 between the baseline and post-dose visits.

#### Prediction of drinking change

Each baseline measure, including operation span (35 measures), was entered into a rank-based regression of drinking change on the measure, arm, and baseline drinking, for drinking days and heavy-drinking days (70 tests, FDR-corrected). The same 35 baseline measures were tested as predictors of PHQ-9 change, adjusting for arm and baseline PHQ-9 (35 tests, FDR-corrected). Robustness checks for the strongest predictor included Pearson models, assessment of the 90-day outcome, leave-one-out samples, adjustment for age, sex, baseline depression, and antidepressant use, exclusion of baseline-only participants, and a processing-accuracy criterion of 85% on the operation span math component.

#### Willingness to exert control

PRISM’s predictions were examined with demand avoidance as the single index. Demand avoidance is not one of the 34 retest measures and was therefore not part of their FDR correction. We tested the drug effect at post-dose testing (ANCOVA); the relation of baseline avoidance to performance (operation span and every performance measure of the same task) and to depressive symptoms; the coupling of change in avoidance with change in drinking and PHQ; and whether baseline avoidance, alone or in interaction with arm, predicted drinking or PHQ change (rank-based regression adjusted for the baseline outcome). These analyses were specified after data collection and are exploratory.

#### Effect sizes and Bayes factors

Because this is a pilot study, we report effect sizes with 95% confidence intervals for all comparisons and, to quantify evidence for the absence as well as the presence of effects, Bayes factors (BayesFactor package; Morey & Rouder, 2026). Drug effects compared an ANCOVA model including arm with a model including only the baseline score, using default priors (Rouder et al., 2012). Correlations used the default stretched-beta prior (Ly et al., 2016), applied to ranks for Spearman correlations, and rank-based regressions compared models with and without the predictor of interest. BF10 expresses the evidence for an effect and BF01 = 1/BF10 the evidence for its absence. Values between 1/3 and 3 were treated as inconclusive and values above 3 as at least moderate evidence (Lee & Wagenmakers, 2013). Table S7 summarizes effect sizes and Bayes factors for the main claims.

Analyses used R 4.4.1 (lme4, lmerTest, BayesFactor, ggplot2) and Python 3.12 (NumPy, pandas, SciPy). The analysis code, which reproduces every statistic from the trial-level files, is available at https://github.com/zceydas/MDDAUD_cogtasks

## Results

### Sample, task validity and clinical course

Demographic and clinical characteristics at baseline did not differ significantly between participants randomized to placebo and to psilocybin (all p≥.18, Welch t-tests or Fisher’s exact tests; Table 2), although participants in the psilocybin arm included more women, had numerically higher PHQ, and had numerically lower QIDS-SR scores.

All five tasks produced the effects they are designed to elicit. Table 3 summarizes these effects and compares them with published values from comparable versions of each task where such values exist.

**Table 3.** Baseline task performance. Expected effects of each task, results in the full baseline sample (demand selection: sessions above chance in the learning phase) and published values from comparable task versions where available. RT, response time.

| Task and measure | Expected effect | Baseline result, mean (SD), t, p | n | Published comparison |
| --- | --- | --- | --- | --- |
| PRL: accuracy | Above-chance learning from gains and losses | Gain .80 (.18); loss .70 (.12); both vs .5, $t(23) \geq 8.16$, $p < 10^{-7}$; gain > loss, $t(23) = 2.88$, $p = .008$ | 24 | Reward .73, punishment .74; no valence difference (healthy adults, 75:25 contingencies; Palminteri et al., 2015) |
| PRL: model fit | Choices predicted by the learning model | Better than random choice in 22 of 24; per-trial likelihood .64 (.09); perseveration median 0.12 (IQR -0.18 to 0.39); inverse temperature median 2.77 (loss trials) and at the upper bound of 10 in 14 of 24 (gain trials) | 24 | Fits of the same model class pooled across two-armed bandit studies (206 participants): choice stickiness centered at 0.15 (SD 1.42); inverse temperature mode 3.4 (Gershman, 2016). No estimates published for this task version (Tichelaar et al., 2023, fixed the learning rate at 0.2) |
| Rule switching: switch cost | Accuracy drops after an unannounced rule change | Learning .82 → .40; transfer .89 → .43; $t(16) = 6.27$ and $9.53$, both $p < 10^{-4}$ | 17 | Same design as the low-volatility condition of Wen et al. (2023), which does not report switch-locked accuracy; that study excluded participants below 65% overall accuracy (6 of 20 here, including the 3 at chance that were excluded) |
| Demand selection: demand effect | Slower, less accurate responses as switch probability rises | RT 0.59 → 0.77 s, $t(18) = 8.81$, $p < 10^{-7}$; accuracy .88 → .82, $t(18) = -3.14$, $p = .006$ | 19 | Linear increase in RT and errors (Sayalı & Badre, 2019, 2021) |
| Demand selection: task-switch cost | Switch trials slower and less accurate than repeat trials | RT 0.19 s (learning), 0.15 s (decision); accuracy 6.6 and 3.6 points; all $t \geq 4.39$, $p \leq .0004$ | 18–19 | Signature effect of cued task switching (Monsell, 2003) |
| Demand selection: demand avoidance | Preference for less demanding decks | 0.54 (0.14), vs .5, $t(17)=1.14$, $p=.27$; no increase with demand difference ($t(17)=0.11$, $p=.91$) | 18 | About half avoiders (Sayalı & Badre, 2019); 67% avoidance (Sayalı & Badre, 2021) |
| MSIT: interference effect | Slower, less accurate interference trials | RT 317 (62) ms, $t(22)=24.56$, $p < 10^{-16}$; accuracy 5.4 points, $t(22)=5.87$, $p < 10^{-5}$ | 23 | 312 (61) ms in healthy adults (Bush et al., 2003) |
| Operation span: recall | Recall declines with set size | Score 9.9 (2.4) of 15 (66% of letters); 86% at set size 3 to 42% at set size 7, $t(16)=-8.60$, $p < 10^{-6}$ | 18 (17) | Partial-credit score 76% of letters (Redick et al., 2012; different scoring) |
| Operation span: processing | Math accuracy $\geq 85\%$ | 87% (7); 11 of 18 meet criterion | 18 | Criterion of Unsworth et al. (2005) |

#### Probabilistic reward learning

Participants chose the better cue on 80% of gain trials (SD 18) and 70% of loss trials (SD 12; both above chance, t(23)=8.16 and 8.36, both p<10⁻⁷, n=24), and learned more accurately from gains than from losses (t(23)=2.88, p=.008). Healthy adults learning comparable 75:25 contingencies choose the better option on about 73–74% of trials, with no difference between reward and punishment (Palminteri et al., 2015; see also Pessiglione et al., 2006). Overall accuracy observed within our sample was in that range, but the advantage for gains over losses is not typical of healthy samples. The reinforcement-learning model predicted choices better than a random-choice model in 22 of 24 participants (likelihood-ratio tests, 5 df, each p<.05; mean per-trial choice likelihood .64). Median learning rates were 0.15 for gains and 0.43 for losses. Inverse temperature reached its upper bound of 10 in 14 of 24 participants on gain trials and 7 of 24 on loss trials, indicating near-deterministic choice once values were learned. Perseveration was small and varied in sign across participants (median 0.12, interquartile range −0.18 to 0.39). Both perseveration and loss-trial inverse temperature were close to the values typical of this model class: pooled across two-armed bandit studies, choice stickiness is centered at 0.15 and inverse temperature has a mode of about 3.4 (Gershman, 2016). Gain-trial inverse temperatures were higher than typical, consistent with near-deterministic choice once reward values had been learned.

#### Probabilistic rule switching

Participants performed above chance in 17 baseline sessions. Accuracy in these sessions was above chance in both the learning phase (.75) and the transfer phase (.76; t(16)=10.26 and 15.50, both p<10⁻⁷) and did not differ between phases (t(16)=−0.96, p=.35). Each unannounced rule change produced the expected drop in accuracy, from .82 before to .40 after a change in the learning phase and from .89 to .43 in the transfer phase (switch costs .43 and .46; t(16)=6.27 and 9.53, both p<10⁻⁴). Post-switch accuracy below .5 indicates that participants initially continued to apply the previous rule before adapting to the new one. Participants did not exceed chance in three of 20 baseline sessions and one of 16 post-dose sessions (one-sided binomial test over 240 trials, p≥.05; accuracy .45–.53), and data from these sessions were excluded (see Data quality). Six of the 20 baseline sessions, including these three, fell below the 65% overall-accuracy criterion that Wen et al. (2023) used for exclusion. Excluding every session below 65% (nine sessions) left the rule-switching results unchanged in direction: 7 of 11 feedback-driven measures moved toward flexibility, as in the primary analysis, and the stability composite still tracked drinking change (*r_s_*=−.54, p=.009).

#### Parametric Demand Selection Task

Participants did not exceed chance in the learning phase in six of 25 baseline sessions, and data from these sessions were excluded (see Data quality). In the remaining 19 baseline sessions, accuracy was .84 in the learning phase and .90 in the decision phase. Raising the task-switch probability from .13 to .88 slowed responses from 0.59 to 0.77 s (t(18)=8.81, p<10⁻⁷) and reduced accuracy from .88 to .82 (t(18)=−3.14, p=.006). This reflects the parametric demand effect that the task is designed to produce (Sayalı & Badre, 2019, 2021). Task-switch costs, the signature effect of cued task switching (Monsell, 2003), were robust in both phases (RT 0.19 s in the learning phase and 0.15 s in the decision phase; accuracy 6.6 and 3.6 percentage points respectively; t(18)=8.60 and t(17)=6.68 for RT, t(18)=5.07 and t(17)=4.39 for accuracy, all p≤.0004). In free choice (during the decision phase), participants were on average close to indifference (demand avoidance .54, SD .14; vs .5, t(17)=1.14, p=.27, n=18): nine participants avoided demand, eight sought it and one was indifferent. This is consistent with performance in the second sample of Sayalı and Badre (2019), which divided almost equally into demand avoiders and demand seekers, and is lower than the 67% avoidance reported by Sayalı and Badre (2021). Unlike Sayalı and Badre (2021), avoidance did not increase with greater difference in demand between the offered decks (.52, .57 and .53 for differences of one, two and three levels, respectively; three vs one level, t(17)=0.11, p=.91), although the probability of choosing a deck fell slightly from the least (.56) to the most demanding deck (.48). Choices therefore discriminated only weakly between demand levels in this sample, which limits the precision of demand avoidance as an index.

#### MSIT

Participants were slower (922 vs 605 ms) and less accurate (.92 vs .97) during interference trials than during control trials (n=23). The interference effect in RT was 317 ms (SD = 62; t(22)=24.56, p<10⁻¹⁶), closely matching the 312 ms (SD = 61) reported in the original validation of the task in healthy adults (Bush et al., 2003). The accuracy cost was 5.4 percentage points (t(22)=5.87, p<10⁻⁵).

#### Operation span

The mean partial-credit score was 9.9 of 15 (SD = 2.4, n=18), equivalent to 66% of letters recalled in the correct position per set. Recall declined with set size, from 86% for three-letter sets to 42% for seven-letter sets (linear decline of 9.5 percentage points per additional letter, t(16)=−8.60, p<10⁻⁶, n=17). Math accuracy averaged 87% (SD 7), and 11 of 18 participants met the conventional 85% criterion (Unsworth et al., 2005). In a large, mostly young normative sample, the partial-credit score averaged 57.4 of 75 letters, or 76% (Redick et al., 2012). That score counts letters rather than set proportions, so the comparison is approximate, but span in this older clinical sample was somewhat lower.

#### Construct validation

At baseline, indices assigned to the same pole shared a small amount of variance across tasks, more than expected by chance (Figure S1A; Table S6). The mean cross-task correlation among aligned indices was *r_s_*=+.11 (permutation p=.032), and 16 of 23 cross-task pairs were positive. The clearest convergences fit the classification. Participants who perseverated more showed smaller MSIT interference reflecting stronger goal shielding (*r_s_*=.57, p=.006, n=22), and larger cued switch costs in accuracy were associated with smaller MSIT interference (*r_s_*=.49, p=.047, n=17). Perseveration was only weakly related to rule-switching cost (*r_s_*=+.21 for the transfer phase, n=16; BF01=1.5, inconclusive). Two features limited coherence. Pre-switch accuracy and the cued RT switch cost tracked general task accuracy (*r_s_*=.53 and −.39 with a composite of overall accuracy across the four tasks, which partly overlaps with these indices by construction), indicating that they index ability as well as stability. Test–retest reliability within arm was modest to low (Figure S1B): *r_s_*=.40 to .45 for MSIT interference, pre-switch accuracy and the cued RT switch cost, and below .30 for the PRL indices and the cued accuracy switch cost. Post-switch accuracy showed no test–retest correlation (*r_s_*=−.01, n=13).

### Overall clinical outcomes

Drinking fell substantially in both allocations (psilocybin and placebo). Over the 30 days after drug administration, the overall proportion of drinking days fell from .62 to .30 and the proportion of heavy-drinking days from .39 to .12 (n=25), with no difference between arms across the baseline, 30-day, and 90-day windows (arm × window, F(2, 45.0)=0.72, p=.49 for drinking days and F(2, 45.0)=0.30, p=.74 for heavy-drinking days; at 30 days, BF01=2.4–2.6, inconclusive). Depressive symptoms fell more after psilocybin (PHQ 9.2 to 2.9) than after placebo (7.5 to 6.1; ANCOVA, d=−0.88 [−1.62, −0.14], t(20)=−2.47, p=.023, BF10=2.7, n=23). Change in PHQ was unrelated to change in drinking (*r_s_*=−.15, p=.48, BF01=1.8).

### Drug effects across the battery

No effect of allocation (psilocybin vs placebo) on task performance survived FDR correction across the 34 analyzed measures (smallest q=.29; Figure 2; Table S1). Effects on the stability–flexibility measures (the first question) are reported in the next section and on the willingness to exert control (the second question) in the section after; this paragraph summarizes the measures outside the continuum. We found no evidence that psilocybin altered inhibition (MSIT interference effect in RT, d=0.20 [95% CI −0.72, 1.12]; accuracy, d=0.35 [−0.47, 1.17]) or value learning (reward-learning accuracy, d=0.16 [−0.79, 1.12]; punishment-learning accuracy, d=0.00 [−0.72, 0.71]). These intervals do not exclude moderate effects, and Bayes factors favored the null hypothesis only weakly (BF01=2.0–2.5). Across all 34 measures, no null result reached even moderate evidence for the absence of a drug effect (largest BF01=2.5; Table S1). Three differences between allocation groups reached uncorrected significance, all in the demand selection task. Two were performance effects: slower learning-phase responding (d=1.01 [0.30, 1.73], t(13)=3.09, p=.009, BF10=7.7) and a larger task-switch cost in RT (d=0.89 [0.07, 1.71], t(13)=2.35, p=.035, BF10=2.4) in the psilocybin arm relative to placebo. The third was a choice effect: a higher baseline-adjusted probability of choosing the least demanding deck in the psilocybin arm relative to placebo (d=1.31 [0.06, 2.55], t(12)=2.29, p=.041, BF10=1.3). The raw means for this measure did not change in the placebo arm (.45 to .44) and fell in the psilocybin arm (.62 to .56). With a baseline imbalance and the strongest baseline–change correlation in the battery (r=−.78), this estimate mainly reflects the baseline adjustment and should not be interpreted as a change in effort choice due to drug administration.

**Figure 1.**
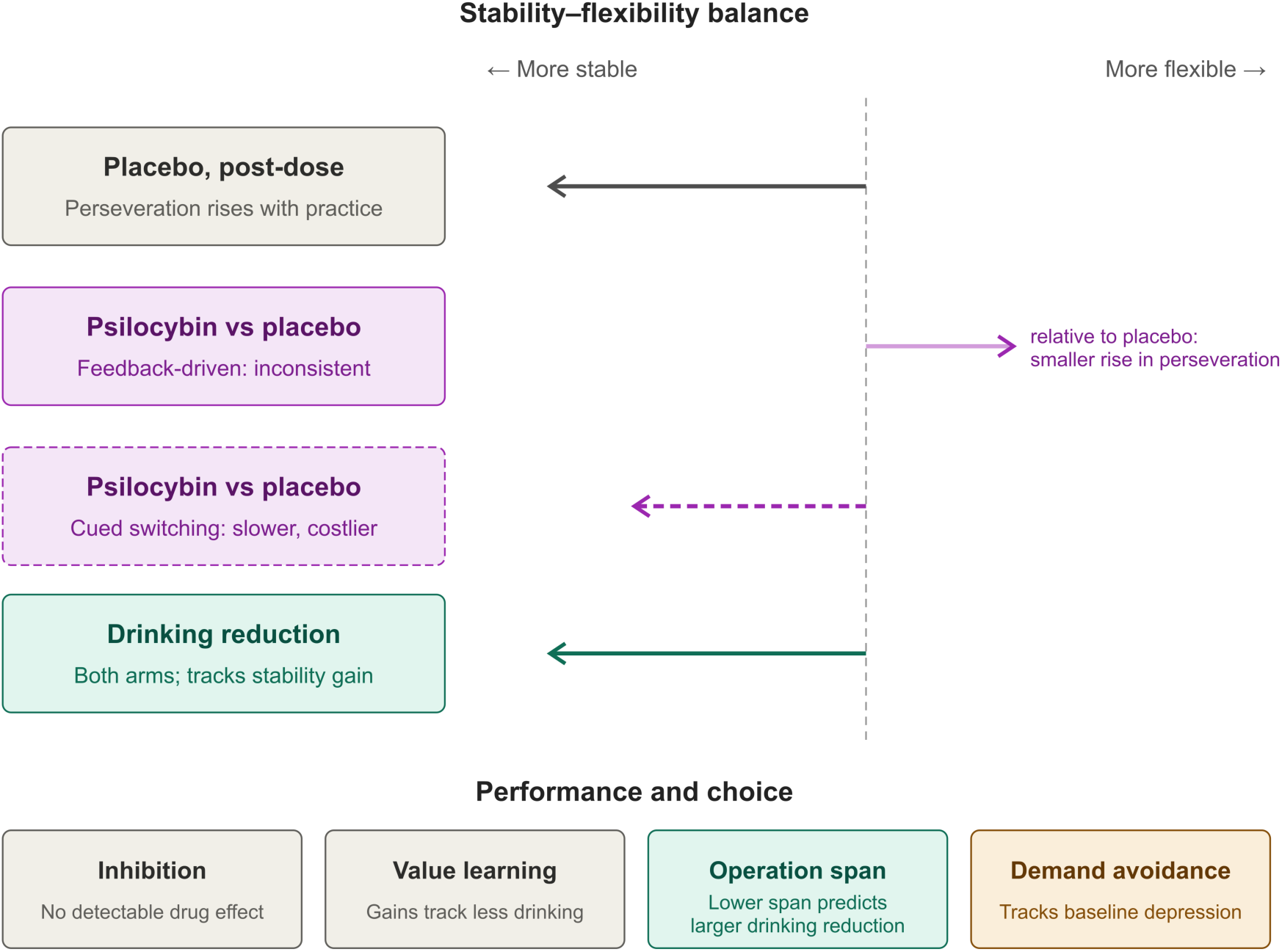
Framework and overview of findings. Top: direction of change along the stability–flexibility continuum. At post-dose testing, perseveration had risen in the placebo arm. Relative to placebo, psilocybin was associated with a smaller rise in perseveration, shown as a relative shift toward flexibility (faint arrow; perseveration did not fall in either arm), although feedback-driven measures as a whole did not move consistently; it also slowed cued task switching (dashed). In both arms, drinking reduction accompanied change toward stability. Bottom: findings for measures outside the continuum. No drug effect was detected on inhibition or value learning; larger increases in reward-learning accuracy accompanied larger drinking reductions; lower baseline operation span predicted larger drinking reduction; demand avoidance, the battery’s measure of willingness to exert control, was unchanged by the drug and tracked depressive symptoms at baseline.

**Figure 2.**
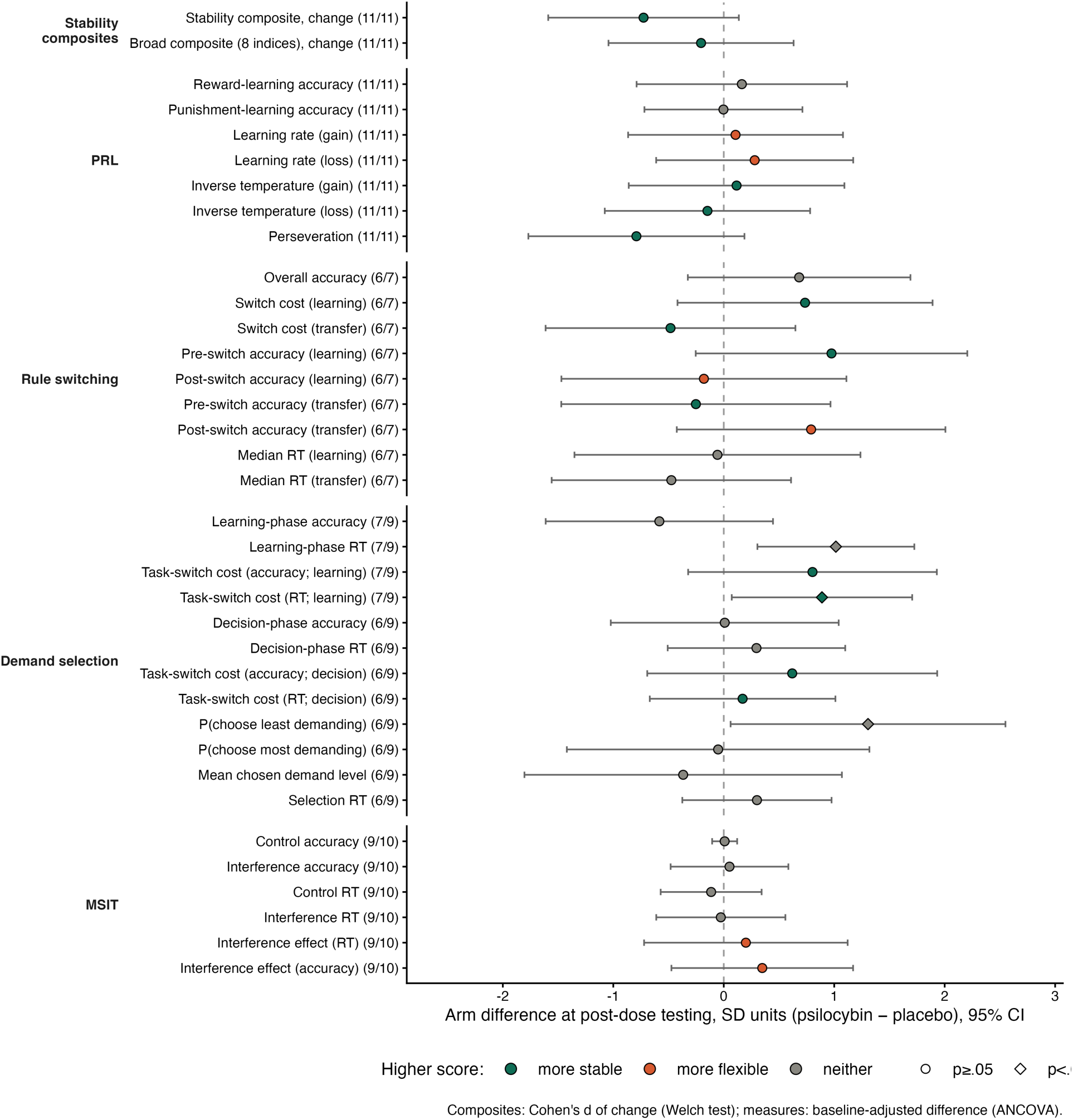
Effect of psilocybin compared to placebo on the stability composites and all 34 retest measures. Baseline-adjusted difference between arms in pooled-SD units (psilocybin minus placebo) with 95% CI; n for placebo/psilocybin in parentheses. Color gives each measure’s position on the stability–flexibility continuum (Table 1). Top rows: the stability composite and the broader eight-index composite (Cohen’s d of change from baseline to post-dose testing; Welch test). Diamonds: uncorrected p<.05; none of the 34 measures survives FDR correction.

### Cued task switching slowed; feedback-driven effects were inconsistent

Re-signing each effect by its position on the continuum showed no consistent direction on the feedback-driven tasks (Figure 3A). On the 11 stability–flexibility measures from PRL and rule switching, psilocybin moved performance toward flexibility relative to placebo in 7 cases (sign test p=.55). The largest effects pointed in different directions: toward flexibility for perseveration (d=−0.79 [−1.77, 0.19], t(19)=−1.69, p=.11) and transfer-phase post-switch accuracy (d=+0.79 [−0.43, 2.01], t(10)=1.45, p=.18), and toward stability for learning-phase pre-switch accuracy (d=+0.98 [−0.25, 2.20], t(10)=1.77, p=.11), all with inconclusive Bayes factors (BF10=0.7–1.2). For perseveration, the difference arose because placebo participants became more stable at post-dose testing, whereas psilocybin participants changed less (Figure 4A): perseveration rose under placebo (0.22 to 0.99, t(10)=3.04, p=.012, n=11) but rose less under psilocybin (0.14 to 0.45, t(10)=1.79, p=.10, n=11). Transfer switch cost showed a similar pattern (d=−0.48 [−1.61, 0.65], t(10)=−0.95, p=.37): it rose under placebo (.35 to .48, t(5)=2.99, p=.031, n=6) but not under psilocybin (.57 to .52, t(6)=−0.66, p=.54, n=7; Figure 4B). No measure moved significantly toward flexibility within the psilocybin arm. The stability composite moved toward stability in the placebo arm (mean z=+0.23, n=11) and toward flexibility in the psilocybin arm (−0.36, n=11), but the difference was not significant (Welch t(19.9)=1.70, p=.10, BF10=1.0; Welch t(10.9)=2.78, p=.018 among the 13 participants with both measures; baseline-adjusted composite, Welch t(19.7)=1.43, p=.17). The broader eight-index composite showed no difference between allocation groups (Welch t(17.7)=0.48, p=.64, BF01=2.4). When including the four rule-switching sessions in which participants performed at chance, 8 of 11 feedback-driven measures moved toward flexibility (sign test p=.23), and the composite difference was larger (Welch t(20.7)=1.96, p=.063).

**Figure 3.**
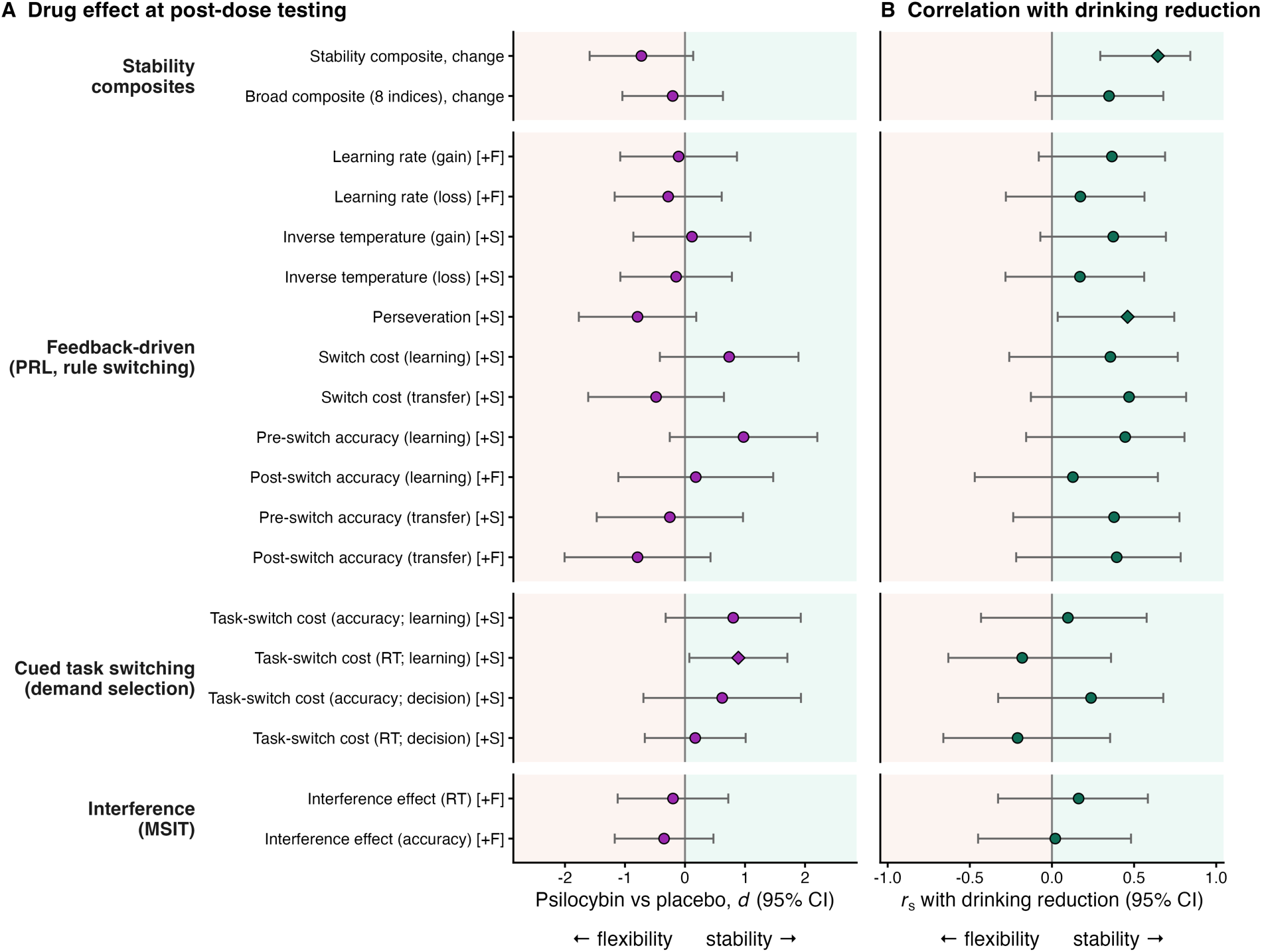
Outcomes organized on the stability–flexibility continuum. [+S] and [+F]: higher scores indicate more stable or more flexible performance (Table 1). Top rows: the stability composite and the broader eight-index composite. (A) Drug effect (psilocybin minus placebo, d with 95% CI; Cohen’s d of change for the composites), re-signed so that positive values indicate movement toward stability. (B) Correlation between change in each measure and change in the proportion of drinking days (Spearman r_s_ with approximate 95% CI), re-signed so that positive values indicate that change toward stability accompanied drinking reduction. Diamonds: uncorrected p<.05; none of the 34 measures survives FDR correction.

**Figure 4.**
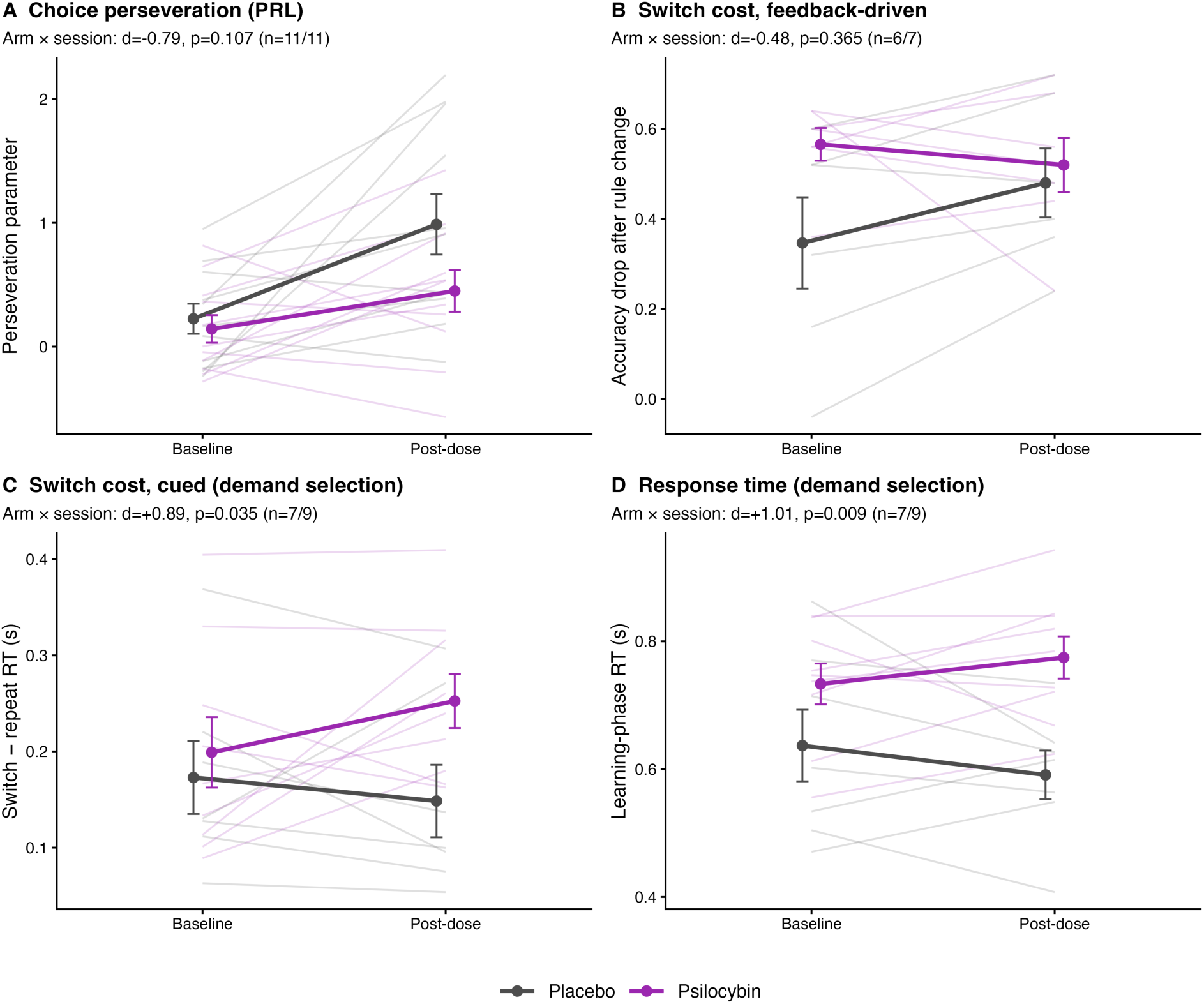
Change from baseline to post-dose testing by arm. (A) PRL perseveration and (B) rule-switching switch cost (transfer phase), two feedback-driven stability measures that rose under placebo and less, or not at all, under psilocybin; neither arm difference was significant. (C) Cued task-switch RT cost and (D) mean RT in the learning phase of the demand selection task, which rose under psilocybin. Thin lines, individuals; thick lines, arm means ± SEM. Statistics from baseline-adjusted ANCOVA.

Cued task switching showed a consistent pattern. All four task-switch costs in the demand selection task moved toward stability under psilocybin relative to placebo (accuracy and RT, learning and decision phases; d=0.17 to 0.89). The learning-phase RT cost rose after psilocybin (0.199 to 0.253 s) and fell after placebo (0.173 to 0.148 s; d=0.89, t(13)=2.35, p=.035; Figure 4C). RT increased overall in the psilocybin arm and decreased in the placebo arm (0.733 to 0.775 s vs 0.637 to 0.591 s; d=1.01, t(13)=3.09, p=.009, BF10=7.7; Figure 4D). Increases in RT were largest on switch trials (d=1.12 [0.46, 1.77], t(13)=3.69, p=.003, BF10=17) and smaller on repeat trials (d=0.64 [−0.08, 1.36], t(13)=1.92, p=.078, BF10=1.5). The slower responses were not accompanied by higher accuracy (learning-phase accuracy d=−0.58, t(13)=−1.22, p=.24), so the slowing did not reflect a shift toward accuracy at the expense of speed. Participants who received psilocybin were slower at baseline; regression to the mean would predict a larger reduction in RT in that arm, but the opposite was observed. After the exclusion of chance-level sessions, these analyses rest on 7 participants who received placebo and 9 participants who received psilocybin. Adjusting for the number of days between drug administration and post-dose testing (median 7 in each arm) did not change these effects (learning-phase RT, t(12)=3.34, p=.006; task-switch cost in RT, t(12)=2.25, p=.044). In mixed models across the four demand levels, the arm × session interaction for learning-phase RT pointed the same way but was weaker (F(1, 16.0)=3.38, p=.084), and the drug did not change how RT, accuracy or choice varied with demand level (arm × level × session, all p≥.21; Table S9).

### No evidence that psilocybin changed the willingness to exert control

At baseline, participants were close to indifference between less and more demanding decks (mean demand avoidance .54, where .5 indicates indifference; t(17)=1.14, p=.27, BF01=2.3, n=18), with wide individual differences. Baseline demand avoidance showed no detectable relation to performance: it was unrelated to operation span (*r_s_*=+.10, p=.75, BF01=1.7, n=13; Figure 5B) and to accuracy, RT and switch costs in the same task (all |*r_s_*|≤.36, all p≥.14, BF01=0.9–2.0, n=18), although with these sample sizes moderate correlations cannot be excluded (95% CI for operation span −.48 to .62). Baseline demand avoidance was not significantly related to stability measures from the other tasks (largest *r_s_*=+.48, with rule-switching cost, p=.12, n=12). It was, however, strongly related to depressive symptoms measured with the PHQ at the same visit: participants with more severe depressive symptoms (higher PHQ-9 scores) avoided demand more (*r_s_*=+.71 [.30, .90], p=.001, BF10=24, n=17; Figure 5A; *r_s_*=+.68, p<.001, n=22 including chance-level sessions). This association was specific to that visit. Baseline demand avoidance was unrelated to depressive symptoms measured at screening with a different instrument, the QIDS-SR (*r_s_*=+.06, n=18), and demand avoidance at post-dose testing was unrelated to PHQ at that visit (*r_s_*=−.16, p=.51, BF01=1.8, n=20). The test–retest reliability of demand avoidance was low (*r_s_*=−.27 after removing arm means, n=15; −.13, n=20 including chance-level sessions), so the baseline association may not generalize beyond that session.

**Figure 5.**
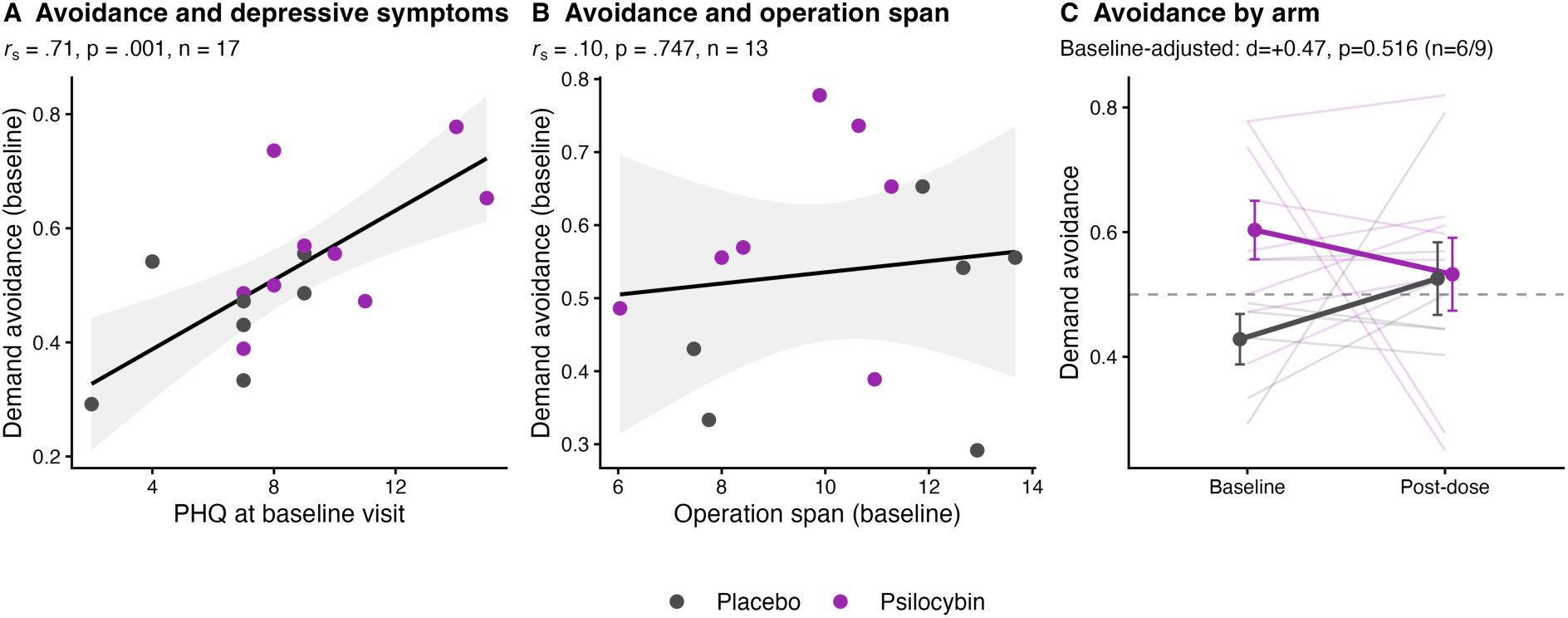
Willingness to exert control. (A) Baseline demand avoidance (proportion of choices of the less demanding deck) was significantly correlated with PHQ at the same visit. (B) Baseline demand avoidance did not significantly correlate with operation span. (C) Demand avoidance at baseline and post-dose testing is plotted separately by allocation; dashed line indicates the point of indifference. Participants whose performance during baseline sessions was at chance in the learning phase are excluded.

There was no evidence that psilocybin increased the willingness to exert control (Table S5). Baseline-adjusted demand avoidance at post-dose testing was, if anything, higher after psilocybin than after placebo (d=+0.47 [−1.06, 2.00], t(12)=0.67, p=.52, n=6/9; Figure 5C), but the Bayes factor was inconclusive (BF01=2.0), so the data neither support nor rule out a drug effect of moderate size. Participants in the psilocybin arm were more avoidant at baseline than those who received placebo (.60 vs .43) and participants in both arms regressed to near .53 after treatment, providing evidence that the estimate is dominated by regression to the mean (baseline–change r=−.77). Change in demand avoidance was unrelated to change in drinking (*r_s_*=−.02, p=.94, BF01=1.9, n=15) and to change in PHQ (*r_s_*=+.27, p=.33, BF01=1.3). Adjusting for treatment allocation and baseline measures on the given outcome, baseline avoidance predicted neither drinking change (rank-partial β=−0.08, t(14)=−0.33, p=.74 for drinking days; β=−0.32, t(14)=−1.48, p=.16 for heavy-drinking days; BF01=2.3 and 2.0) nor PHQ change (β=+0.08, t(13)=0.64, p=.54), and it did not interact with allocation (interaction |t|≤0.54, p=.60–.75). For PHQ change, the Bayes factors provided moderate evidence against both the main effect of baseline avoidance (BF01=3.4) and the interaction of avoidance with allocation (BF01=3.3). Without adjustment, more avoidant participants showed larger PHQ reductions (*r_s_*=−.61, p=.009, n=17), an association explained by their higher baseline PHQ. Within the psilocybin arm, more avoidant participants showed somewhat larger PHQ reductions (unadjusted *r_s_*=−.50, p=.14, n=10).

### Change toward stability accompanied drinking reduction

No association between cognitive change and drinking change survived FDR correction across the 34 measures (smallest q=.49; Table S2). Here and below, drinking change is the change in the proportion of drinking days from baseline to the 30 days after drug administration. The sign of the correlations was nevertheless consistent. On 15 of the 17 stability–flexibility measures, change toward stability was associated with larger drinking reductions (descriptive sign test, 15 of 17, p=.002; Figure 3B). This held for all 11 feedback-driven measures and for 2 of the 4 cued switch costs. The strongest correlations with drinking change were for reward-learning accuracy (*r_s_*=−.48, p=.022, BF10=3.7, n=22; adjusted for arm, t(19)=−2.27, p=.035; Figure 6B) and perseveration (*r_s_*=−.46, p=.031, BF10=2.9, n=22; adjusted for arm, t(19)=−2.11, p=.048). Among the rule-switching measures, the largest correlation, for transfer-phase switch cost, was similar in size (*r_s_*=−.47) but less certain, because only 13 participants contributed (p=.11). The stability composite correlated more strongly with drinking change than any single measure (*r_s_*=−.64, p=.001, BF10=29, n=22; adjusted for arm, t(19)=−3.31, p=.004, BF10=16; additionally adjusted for the interval between drug administration and post-dose testing, t(18)=−3.09, p=.006; Figure 6A). This correlation did not differ by arm (arm × composite interaction, t(18)=−0.47, p=.64) and was present in each arm (placebo *r_s_*=−.45, psilocybin *r_s_*=−.60). Among the 13 participants with both component measures, the correlation between composite change and drinking change was weaker and less certain (*r_s_*=−.56 [−.86, .04], p=.049, BF10=2.2; adjusted for arm, t(10)=−1.08, p=.31). For the other nine participants the composite consists of perseveration change alone, so part of the composite’s correlation reflects the correlation of perseveration change with drinking change. When change was adjusted for baseline scores, the composite’s correlation with drinking change was weaker (*r_s_*=−.43 [−.73, .01], p=.044, n=22; adjusted for arm, t(19)=−1.79, p=.090), so part of the association reflects differences in where participants started. The broader eight-index composite showed a weaker correlation in the same direction (*r_s_*=−.35, p=.11, BF10=1.3). Concurrently, higher perseveration at post-dose testing was associated with fewer heavy-drinking days over the 30 days after drug administration (*r_s_*=−.51, p=.014; adjusted for arm and baseline perseveration, t(18)=−2.97, p=.008; Figure 6C).

**Figure 6.**
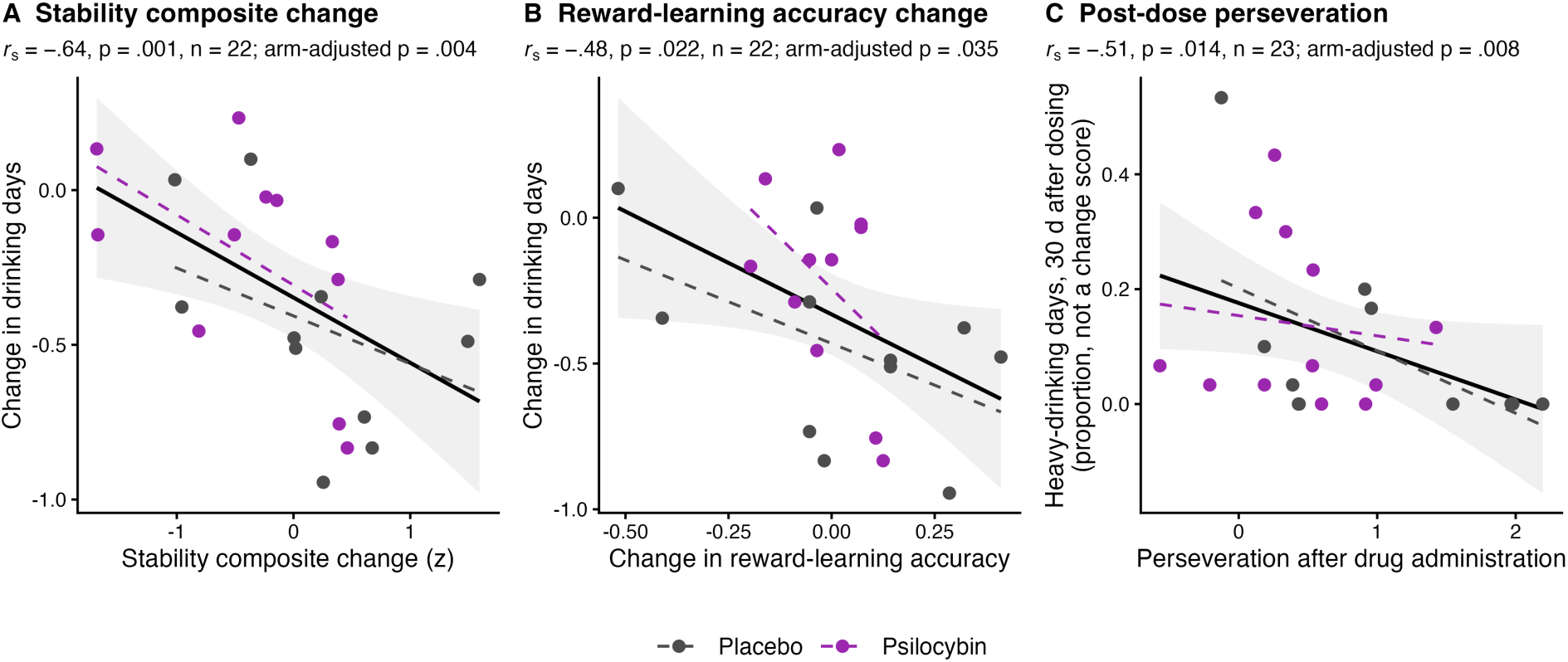
Pre-post changes in stability and reward learning were associated with drinking reduction in both experimental groups. (A) Change in the stability composite and (B) change in reward-learning *accuracy are plotted against change in the proportion of drinking days from baseline to 30 days after drug administration. (C) Perseveration after drug administration is plotted against the proportion of heavy-drinking days over the 30 days after drug administration (a level, not a change score; see text); the adjusted p also controls for baseline perseveration. Black lines depict pooled line of best fit, with shaded areas representing 95% CI; dashed lines depict fit within each allocation*.

### Lower working-memory span predicted larger drinking reduction

Of 70 prognostic tests (35 baseline measures × 2 drinking outcomes), baseline operation span was the strongest predictor. Greater span predicted smaller reductions in heavy-drinking days (rank-partial β=+0.57, t(14)=3.94, p=.0015, BF10=25) and total drinking days (β=+0.63, t(14)=3.76, p=.0021, BF10=19), adjusting for arm and baseline drinking (n=18; q=.074 for both; Figure 7). The next strongest predictor showed the same direction of association: higher decision-phase accuracy in the demand selection task predicted smaller reductions in drinking days (β=+0.53, t(14)=3.28, p=.0055, q=.13).

**Figure 7.**
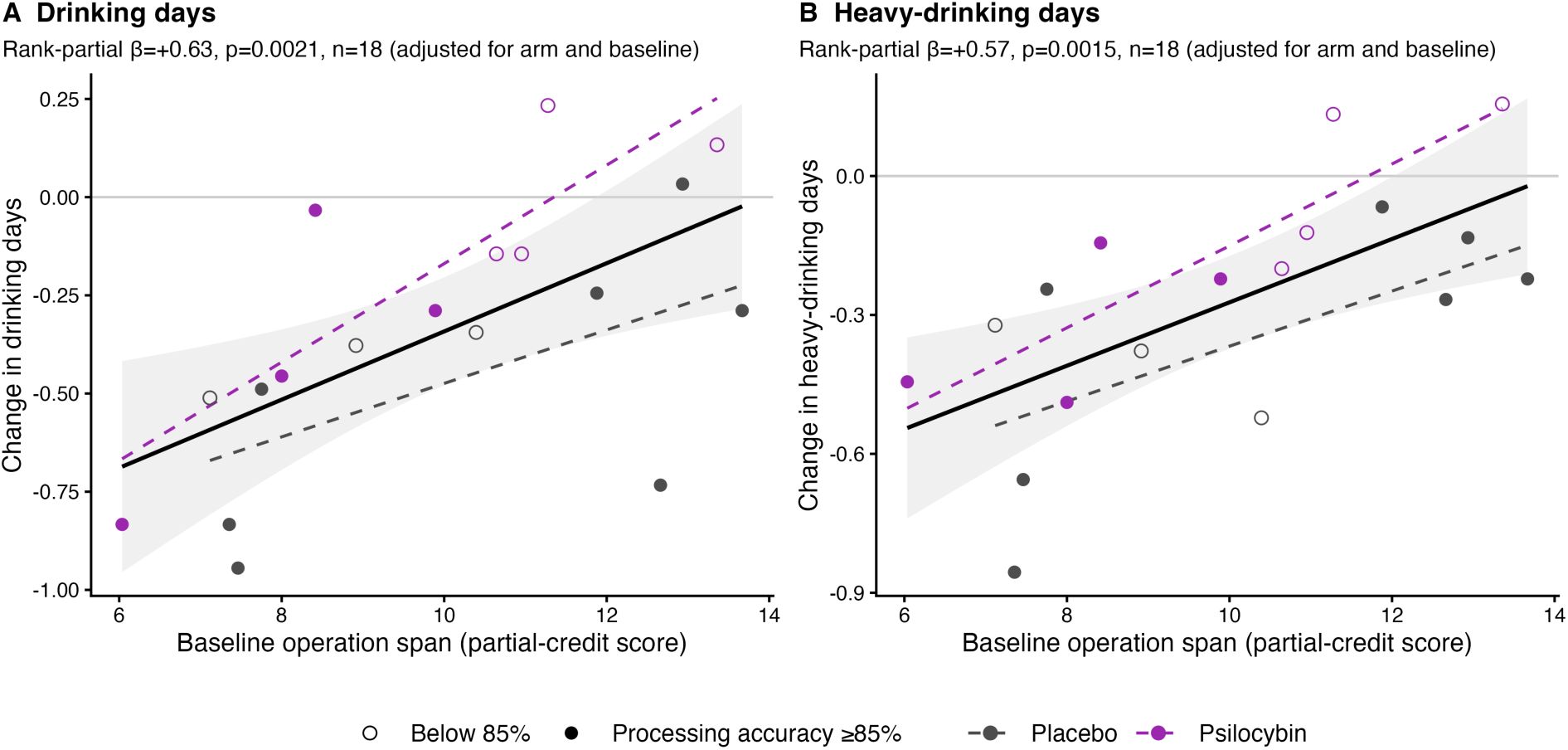
Lower baseline operation span predicts larger drinking reduction. Change from baseline to the 30 days after drug administration in (A) the proportion of drinking days and (B) the proportion of heavy-drinking days are plotted against baseline operation span (n=18; placebo 10, psilocybin 8). Open symbols: processing (math) accuracy below 85% at baseline. Rank-based partial regression adjusted for allocation and baseline drinking. Black lines depict pooled line of best fit, with shaded areas representing 95% CI; dashed lines depict fit within each allocation.

The relationship between operation span and drinking change was present in both arms (placebo *r_s_*=.67–.70, n=10; psilocybin *r_s_*=.83–.90, n=8) and did not differ significantly between them (arm × span interaction, t(13)=1.61, p=.13 for heavy-drinking days and t(13)=1.03, p=.32 for drinking days; BF01=1.3–1.9). It held for the 90-day window (t(13)=4.00, p=.0015, and t(13)=4.27, p=.0009, for heavy-drinking and drinking days), in linear (Pearson) models (t(14)=3.95, p=.0015, and t(14)=2.84, p=.013), in every leave-one-out sample (all t≥3.00, largest p=.010), without the baseline-only participant who had a span score (n=17; t(13)=3.82, p=.0021, and t(13)=3.69, p=.0027) and after adjustment for age, sex, depressive symptoms at screening or antidepressant use (t(13)=2.27–4.26, p=.0009–.041; Table S4). Among the 11 participants who met the 85% processing-accuracy criterion, the estimates remained positive but were less precise: β=+0.56 for heavy-drinking days (t(7)=1.99, p=.087) and β=+0.83 for drinking days (t(7)=2.97, p=.021). Participants with lower span tended to drink more often at baseline (*r_s_*=−.42). Lower span was also associated with larger increases in reward-learning accuracy after drug administration (*r_s_*=−.63, p=.006, n=17), but not detectably with change in rule-switching cost (*r_s_*=−.18, n=12).

### Change in depressive symptoms was not associated with cognitive change

We applied the same analyses to depressive symptoms. Change in PHQ-9 between the baseline and post-dose visits was not associated with change in any cognitive measure (no pooled correlation reached p<.05; smallest q=.94; n=13–22). After adjustment for arm, one association reached uncorrected significance: larger increases in the decision-phase RT switch cost accompanied larger PHQ-9 reductions (rank-partial β=−0.37, t(12)=−2.98, p=.011, n=15; Table S2), but it did not survive correction (q=.39). Unlike drinking, PHQ-9 change showed no consistent direction on the continuum: change toward stability accompanied PHQ-9 reduction on 11 of 17 measures (sign test p=.33), including 6 of the 11 feedback-driven measures, compared with 15 of 17 for drinking reduction. Change in PHQ-9 was also unrelated to change in the stability composite (*r_s_*=+.04, p=.85; adjusted for arm, t(19)=−0.88, p=.39; BF01=2.2) and in drinking (*r_s_*=−.15, p=.48). None of the 35 baseline performance measures predicted PHQ-9 change after adjustment for arm and baseline PHQ-9 (smallest q=.24). In particular, operation span, the strongest predictor of drinking reduction, did not predict PHQ-9 change (rank-partial β=−0.04, t(13)=−0.20, p=.84, BF01=2.9, n=17). The battery’s one correlate of depressive symptoms was a baseline level of a choice measure, not a performance measure or a change: baseline demand avoidance tracked PHQ-9 at the same visit (*r_s_*=+.71, BF10=24; see above). This did not extend to change. Change in avoidance was unrelated to change in PHQ-9 (*r_s_*=+.27, p=.33), and baseline avoidance did not predict PHQ-9 change once baseline PHQ-9 was taken into account (β=+0.08, p=.54, BF01=3.4). Because PHQ-9 was measured at the two cognitive visits, these are associations between concurrent changes over about one week, whereas drinking change was measured over the 30 days after drug administration.

### Associations between baseline measures and baseline drinking behavior

No baseline association between cognition and drinking survived correction across 105 comparisons (smallest q=.41; Table S3). Participants who perseverated more at baseline drank on fewer days but drank more on the days they drank (drinking days *r_s_*=−.42, p=.043; drinks per drinking day *r_s_*=+.57, p=.004). A smaller MSIT interference effect showed the same association with drinks per drinking day (*r_s_*=−.55, p=.008) but not with drinking days (*r_s_*=+.12, p=.58). Perseveration and the MSIT interference effect were themselves correlated (*r_s_*=−.57, p=.006). Participants who perseverated more and were less affected by interference at baseline therefore tended to drink less often but more heavily.

## Discussion

We examined two cognitive-control accounts of the effects of psilocybin in patients with co-occurring AUD and depression, using every measure a five-task battery produced. In this pilot study, no single drug effect or clinical coupling survived correction across the battery, so our conclusions rest on effect sizes, on the consistency of direction across measures, and on the few effects with at least moderate Bayesian support. Within those limits, four findings emerged. Approximately one week after dosing, psilocybin slowed cued task switching relative to placebo, whereas its effects on feedback-driven measures were inconsistent in direction; the largest was on perseveration, which rose less with practice under psilocybin than under placebo. We found no evidence of change in inhibition, value learning, or the willingness to exert control, although Bayes factors showed that these null results are inconclusive rather than evidence of absence. Drinking fell in both arms, and on nearly every stability–flexibility measure the change that accompanied drinking reduction was toward stability, not flexibility; no such pattern was found for depressive symptoms. Lower baseline working-memory span predicted larger drinking reductions in either arm.

### Placing the battery on the stability–flexibility continuum

Figure 3 shows, for every measure with a position on the continuum, both the drug effect and the coupling with drinking.

#### Probabilistic reward learning

Perseveration is the clearest stability index in the battery, because it captures the tendency to repeat a choice regardless of its outcome. It rose at post-dose testing under placebo and less under psilocybin, and larger increases were associated with larger drinking reductions. Inverse temperature, the exploitation of learned values, showed no consistent drug effect, but change in inverse temperature was associated with change in drinking in the same direction as perseveration. Learning rates, the flexibility side of value learning, showed small drug effects toward flexibility; however, the association between change in drinking rates and change in learning rates pointed toward stability, in that faster updating was associated with smaller drinking reductions.

#### Rule switching

Decomposing the switch cost was informative, although with 6 placebo and 7 psilocybin participants the estimates are imprecise. The drug effect on switch cost came mainly from an effect of drug on post-switch accuracy, observed as faster adaptation to a new rule (transfer phase d=+0.79), whereas pre-switch accuracy, which reflects maintenance of the current rule, showed effects in opposite directions in the two phases (d=+0.98 in the learning phase and −0.25 in the transfer phase). In their coupling with drinking, both components pointed toward stability: better rule maintenance and slower adaptation were associated with drinking reduction, although no single rule-switching coupling was significant (the largest was for transfer-phase switch cost, *r_s_*=−.47, p=.11).

#### Cued task switching

All four task-switch costs moved toward stability under psilocybin, in contrast to the mixed feedback-driven measures, and their coupling with drinking was mixed (2 of 4 pointed toward stability, and none was significant). Changes in cued switch costs were unrelated to changes in rule-switching costs (*r_s_*=−.02, n=9), so the two tasks do not appear to index a single shifting ability.

#### MSIT

Interference effects showed no drug effect and weak couplings in the stability direction. We placed them at the flexibility pole as an index of weaker goal shielding. Change across tasks was consistent with this placement, although not significantly: as rule-switching costs rose, MSIT interference tended to fall (*r_s_*=−.49, p=.13, n=11), so greater stability in one task tended to be associated with stronger goal shielding in the other.

#### Integrating findings across tasks in the battery

Two conclusions follow. First, the classification captured a modest amount of variance common to measures from different tasks: participants who were more stable on one task tended, weakly, to be more stable on others. At baseline, indices assigned to the same pole correlated weakly across tasks (mean *r_s_*=+.11, p=.032), with the clearest convergence between perseveration and goal shielding in the MSIT (Figure S1). Limited test–retest reliability, a common property of behavioral control measures (Hedge et al., 2018), and ability-related variance in accuracy-based indices constrain how coherent stability and flexibility can appear as traits in a battery of this size; perseveration and rule-switching cost, for example, were only weakly correlated (*r_s_*=+.21). The contrast between the consistent drug effect on cued switching and the mixed drug effects on feedback-driven measures is therefore better interpreted as a difference between forms of flexibility than as evidence against the classification. For the same reason, the primary stability composite combined only the two feedback-driven stability measures; the broader composite, which also included cued switch costs and MSIT interference, showed weaker effects. Second, associations with drinking were strikingly uniform across feedback-driven measures: more stable performance at post-dose testing was associated with less drinking.

### Feedback-driven and cued flexibility after psilocybin

Sayalı et al. (2026) predicted a selective post-acute gain in flexibility, most clearly in feedback-driven set shifting of the kind measured by card-sorting tasks, with reduced perseveration and no change in inhibition. Our data offer at most weak support for this prediction. We found no evidence of change in inhibition, but feedback-driven measures did not move consistently toward flexibility relative to placebo (7 of 11). The largest effect was on perseveration, the measure closest to the perseverative errors previously found to reduce after treatment with psilocybin (Doss et al., 2021), and this effect took the form of a smaller practice-related rise rather than a treatment-related gain: perseveration rose at post-dose testing under placebo and less under psilocybin, and no measure improved above baseline within the psilocybin arm. This estimate (d=−0.79) was imprecise and its Bayes factor inconclusive. The overall pattern also depended on four rule-switching sessions in which accuracy did not exceed chance: with these sessions included, 8 of 11 feedback-driven measures moved toward flexibility, but such sessions cannot index rule-based control. A larger trial with a placebo arm is needed to establish whether psilocybin prevents the practice-related stabilization of feedback-driven choice, an effect that a design without a placebo arm would miss.

Cued task switching showed the clearest drug effect, in a form of flexibility whose response to psychedelics had not been systematically studied (Sayalı et al., 2026): switching between instructed task sets became slower and costlier. The two forms of shifting, explicitly cued and feedback-inferred, differ in what signals the switch. In feedback-inferred switching (the rule-switching task), the need to switch must be inferred from probabilistic feedback, so performance depends on how readily current commitments are relaxed. In explicitly cued switching (the demand selection task), the switch is instructed and performance depends on how efficiently a task set is reconfigured. Meta-control accounts posit that affective and motivational states shift the balance between stability and flexibility (Goschke, 2003), and positive affect, for example, reduces perseveration at the cost of greater distractibility (Dreisbach & Goschke, 2004). A loosening of learned, value-based commitments after psilocybin would predict reduced perseveration, the direction of the largest feedback-driven effect. We did not, however, observe the distractibility cost that such accounts predict, as MSIT interference was unchanged, although our power to detect it was limited.

Two differences may explain why we did not observe the reduced perseveration that Doss et al. (2021) found one week and one month after treatment. The more likely is the treatment and the population: their participants received two doses of psilocybin rather than one, and had MDD without co-occurring AUD. Timing is a second possibility. Our post-dose session (4–9 days after a single dose) overlapped their one-week assessment, but that assessment followed a second dose, and Sayalı et al. (2026) note that early post-acute effects can diverge from later ones: one day after a low dose of LSD, cognitive flexibility was still impaired while verbal fluency had improved (Wiessner et al., 2022), and in the pig brain, serotonin 2A receptor density was reduced one day after psilocybin and back at baseline one week later (Raval et al., 2021). Whether the pattern we observed changes at later time points requires repeated testing.

### What performance changes can and cannot reveal

Because performance reflects both ability and the control a person chooses to invest, the slowing of cued task switching has more than one explanation. It may reflect less efficient reconfiguration of task sets, or a change in capacity. Alternatively, it may reflect less intense engagement within the task: reduced response vigour, of the kind expected when the opportunity cost of time changes (Niv et al., 2007; Kurzban et al., 2013; Otto & Daw, 2019). Or it may reflect general psychomotor slowing, which in acute studies affects overall response time more than process-specific executive measures (Yousefi et al., 2025). Three features of the current data constrain these explanations without selecting between them. Slowing was largest on switch trials but remained present on repeat trials; accuracy did not improve as slowing increased; and free choice between demand levels did not shift toward easier decks. If reduced engagement is the explanation, it likely operated on the intensity of control within trials rather than on the decision to take on demanding work. The baseline-adjusted increase in choices of the least demanding deck is consistent with this, but the baseline imbalance in choice behavior and raw means make it difficult to confidently support this interpretation.

The same caveat applies to every performance measure in the battery, including the feedback-driven effects and operation span. Distinguishing motivation from other aspects of performance requires measures that vary the value of control directly, such as incentive manipulations, effort-discounting choices (Westbrook et al., 2013), or ratings of effort and task salience collected alongside performance. Thus, one cannot infer motivation from performance after the fact (Sayalı, 2026).

### The willingness to exert control

The demand selection task provided the battery’s one direct measure of the willingness to exert control.

First, willingness to exert control and performance were separable at baseline. Demand avoidance showed no detectable relation to operation span or to performance in the task being chosen, consistent with earlier evidence that avoidant participants are not less able (Sayalı & Badre, 2019). With 13 to 18 participants, this is an absence of evidence rather than proof of independence (BF01=0.9–2.0). Avoidance, however, was strongly related to depressive symptoms at the same visit (BF10=24), matching the profile of behavior that is predicted by motivational accounts of depression (Grahek et al., 2019). Two observations temper this result. The association was not present at post-dose testing, when many participants’ symptoms had fallen, and test–retest stability of avoidance was low. Avoidance in this sample therefore behaved either as a state measure or as a measure with limited reliability over repeated administration; Sayalı (2026) cites evidence that avoidance on this task tracks trait need for cognition. A stable trait would predict higher test–retest reliability than we observed, so the low reliability here more likely reflects measurement noise in a small sample than a change in the trait; a larger sample could test this directly. The baseline association was exploratory and needs replication.

Second, we found no evidence that psilocybin increased willingness to exert control about one week after dosing, although it reduced depressive symptoms more than placebo. Sayalı (2026) identifies this pattern (no increase in willingness to engage effortful tasks on a demand-selection paradigm despite symptom improvement) as evidence that would challenge the model’s core hypothesis. The model also states that increased engagement is expected for personally meaningful goals and that increased engagement with tasks lacking personal meaning would contradict it. Our task was neutral, so this boundary condition could explain the null result. Invoking it after the fact, however, would be the kind of post hoc reasoning Sayalı (2026) warns against, particularly as the model itself proposes this neutral paradigm as its test. Our data therefore provide no support for a general post-acute increase in the willingness to exert cognitive effort about one week after dosing, but they are not strong evidence against such a change in willingness: the sample for comparison between allocations was small (6 vs 9), estimated confidence intervals were wide (d from −1.06 to 2.00), and the estimated Bayes factor for the comparison between allocations was inconclusive (BF01=2.0). The personal meaningfulness of goals was not measured at either session. The prediction therefore remains open for a larger trial that offers both meaningful and neutral demanding choices.

Third, the prediction that participants with high effort avoidance and preserved performance would benefit most was not supported: in models adjusted for arm and for the baseline value of each outcome, baseline avoidance predicted neither drinking nor PHQ change, alone or in interaction with allocation, and for PHQ change the evidence against the prediction was moderate (BF01=3.3–3.4). Without this adjustment, more avoidant participants in the psilocybin arm showed larger PHQ reductions (*r_s_*=−.50, p=.14), the direction the prediction expects. However, more avoidant participants also had higher baseline PHQ scores, and participants with higher baseline scores had more room to improve; once baseline PHQ was taken into account, baseline avoidance no longer predicted PHQ change (β=+0.08, p=.54). The unadjusted pattern therefore most likely reflects higher baseline symptoms rather than a benefit of avoidance.

### Drinking reduction and cognitive stability

If psilocybin reduces drinking by increasing flexibility, increases in flexibility should accompany drinking reductions. Across the battery, the opposite was observed, in both arms. Greater perseveration, larger switch costs, better rule maintenance, greater exploitation, and better reward learning were all associated with larger drinking reductions, and higher perseveration at post-dose testing was associated with fewer concurrent heavy-drinking days. Drinking reduction was thus associated with more stable, consistent, and value-guided choice. This fits accounts in which AUD involves weak goal maintenance and in which reducing drinking requires a newly adopted goal to be shielded from competing cues (Goschke, 2014). The evidence is correlational and rests mainly on the feedback-driven measures; the broader composite, which included cued switch costs and MSIT interference, showed a weaker relationship, and the association with the stability composite was also weaker when change was adjusted for baseline scores. The direction of influence is also unknown. Drinking less may itself make performance more consistent, for example through fewer residual effects of alcohol on testing days.

Depressive symptoms demonstrated a different relationship with cognition. Psilocybin reduced depressive symptoms in this sample more than placebo, although the Bayesian evidence for this was only anecdotal (BF10=2.7). Change in depressive symptoms was unrelated to change in drinking, was not associated with change on any cognitive measure or with the stability composite, showed no consistent direction on the stability–flexibility continuum (11 of 17 measures, against 15 of 17 for drinking), and was not predicted by working-memory span. One possibility is that change in cognitive control is a transdiagnostic mechanism whose beneficial direction differs between disorders: depression is often characterized by rigidity that may benefit from greater flexibility (Sayalı et al., 2026), whereas reduced drinking may require greater stability. Our data do not test this directly, as we did not find evidence that mood change was associated with either pole of the stability-flexibility continuum. At baseline, depressive symptoms were associated with the willingness to exert control rather than with any performance measure, and this is a link that future work could investigate longitudinally. The comparison between the two outcomes is limited by design: depressive symptoms were measured by self-report at the two cognitive visits, about one week apart, whereas drinking change covered 30 days, and the parent trial’s clinician-rated depression outcome was not analyzed here.

### Working-memory span as a predictor of drinking reduction

Lower baseline working-memory span predicted larger drinking reductions in both allocations. It was the strongest of 70 prognostic tests (q=.074; BF10=19–25) and survived every robustness check, and baseline accuracy in the demand selection task yielded the same direction of association with change in drinking behavior. This is contrary to the common expectation that better cognitive function supports better treatment outcomes (Bates et al., 2013). Several explanations are possible. Participants with lower span tended to drink more often at baseline, and although baseline drinking was included as a covariate, residual regression effects cannot be excluded with 18 participants. Extrapharmacological factors may also be relevant: people with lower span may rely more on external structure, so the frequent assessment and therapeutic contact of a trial may benefit them more (Clifford & Maisto, 2000). Lower span was also associated with larger increases in reward-learning accuracy, although not detectably with change in rule-switching cost, which may suggest that participants with the most room to improve changed most. Finally, as a performance measure, span reflects engagement as well as capacity. The effect of baseline working-memory span predicting reductions in drinking held among participants who met the processing-accuracy criterion, and span showed no detectable relation to demand avoidance, which argues against an explanation in terms of effort, but does not exclude it. Because the effect did not depend on allocation, span appears to be a general prognostic marker rather than a marker of response to psilocybin. It did not predict change in depressive symptoms (BF01=2.9), so its prognostic value may be specific to drinking.

### Effect sizes for future trials

Because this is a pilot study, its main contribution is the size and precision of effects rather than tests of significance, and we report standardized effect sizes with confidence intervals and Bayes factors for every comparison (Tables S1 and S7). Four findings were supported by at least moderate evidence: the slowing of cued task switching after psilocybin (switch trials d=1.12, BF10=17; learning-phase RT d=1.01, BF10=7.7), the association between baseline demand avoidance and baseline depressive symptoms (BF10=24), the prediction of drinking reduction by working-memory span (BF10=19–25), and the coupling of drinking reduction with change in the stability composite (BF10=29, weaker after adjustment for baseline scores) and in reward-learning accuracy (BF10=3.7). For depressive symptoms, there was also moderate evidence against the prediction that baseline demand avoidance would forecast improvement (BF01=3.4). No drug effect, by contrast, was supported by even moderate evidence for its absence: Bayes factors for the null did not exceed 2.5, so these results are insensitive rather than negative. Effect sizes from pilot studies are imprecise and should not be used directly for power calculations (Leon et al., 2011). Future trials should instead be powered for the smallest effect of interest, using the confidence intervals reported here as a guide to plausible ranges. On that basis, the results most worth confirming are the slowing of cued switching, the association between change toward stability and drinking reduction, and the prognostic value of working-memory span. The smaller practice-related rise in perseveration under psilocybin than under placebo (d=−0.79, with an interval that includes zero) is a candidate effect rather than a finding.

### Limitations

- The sample was small: 12 to 25 participants per analysis. After the exclusion of chance-level sessions, drug effects rested on 6 placebo and 7 psilocybin participants for rule switching, 7 and 9 for demand selection performance and 6 and 9 for demand avoidance. No single drug effect or coupling survived correction across the battery, and confidence intervals around null effects were wide; Bayes factors showed that these null results are inconclusive rather than evidence of absence.
- The classification of measures followed construct definitions but was finalized after data collection, and some placements are debatable. Inverse temperature conflates exploitation with choice noise, and cued switch costs may reflect reconfiguration efficiency or engagement as much as task-set inertia. The sign tests treat correlated measures as independent and are descriptive. At baseline, indices assigned to the same pole shared only modest variance across tasks, and several had low test–retest reliability (Figure S1).
- The stability composite was defined after data inspection. A broader composite showed weaker effects, and adjusting change for baseline scores weakened the composite’s association with drinking reduction. The composite, the motivation analyses and the clinical-coupling analyses are exploratory.
- Every score except free choice is a performance measure and reflects both ability and the control invested. Willingness to exert control was measured with a single neutral choice task whose test–retest stability was low. Task salience, subjective effort and personal meaning were not measured, so PRISM’s specificity claim could not be tested. Depressive symptoms were measured by self-report (PHQ-9) at the cognitive visits only.

### Conclusions

In this pilot study, about one week after dosing, psilocybin did not uniformly enhance cognitive flexibility, and there was no evidence that it increased the willingness to exert control. Relative to placebo, it slowed cued task switching, had no consistent effect on feedback-driven measures, and showed no detectable effect on inhibition or value learning. Drinking fell in both arms, and its reduction was associated with movement toward stability on nearly every measure and was predicted by lower working-memory span. Depressive symptoms improved more after psilocybin, but their improvement was not associated with cognitive change. A transdiagnostic cognitive-control account of psychedelic therapy may therefore need to specify the form of flexibility involved, to distinguish performance from the willingness to exert control, and to specify the direction of change that recovery requires in each disorder.

## Supporting information

Sayali_psilocybin_cognition_supplement

## Ethics

The current trial (IRB00259934) and the parent trial (IRB00233684) were approved by the Johns Hopkins Medicine Institutional Review Board, and the parent trial was registered at ClinicalTrials.gov (NCT04620759). All participants provided written informed consent.

## Data availability

The data are available from the corresponding author upon reasonable request, subject to the conditions of the trial’s ethics approval.

## Code availability

The analysis code, which reproduces every statistic, table and figure in this article and its supplement from the raw task files, is available at https://github.com/zceydas/MDDAUD_cogtasks

## Author contributions

CS: Conceptualization, Software, Formal Analysis, Writing – original draft. EW: Investigation, Writing – review & editing.

FSB: Conceptualization, Writing – review & editing.

## Acknowledgments

We thank the participants, and the clinical and research team of the parent trial.

