## Supplementary material for "Psilocybin, cognitive stability and drinking reduction in co-occurring alcohol use disorder and depression: a pilot secondary analysis of a randomized, placebo-controlled trial": Sayali_psilocybin_cognition_supplement

#### Supplementary Methods

Scoring from trial-level data. All measures were scored from the trial-level files written by each task. A few files had been saved outside the expected task folder (for example, two baseline operation span files and one baseline rule-switching file inside the reward-learning folder); files were therefore located by searching each session folder, and duplicate copies were resolved explicitly.

Probabilistic reward learning. Missed responses are coded 999 in the response-time and accuracy columns and were removed before computing accuracy and before model fitting. The task's own accuracy column records whether the trial's outcome was favourable rather than whether the better cue was chosen, so learning accuracy was computed from the chosen-cue accuracy column. The Rescorla–Wagner model was fitted by maximum likelihood (L-BFGS-B, best of 50 random starts with a fixed seed). In three sessions a learning rate at its lower bound left the corresponding inverse temperature undetermined; perseveration estimates were unaffected by the choice of starting values.

Probabilistic rule switching. Pre-switch and post-switch accuracy were computed around every change of the relevant rule (three in the learning phase and five in the transfer phase). Feedback was valid on 80% of trials in every session. Sessions whose overall accuracy did not exceed chance (one-sided binomial test over 240 trials, $p \geq .05$; three baseline sessions and one post-dose session) were excluded from all rule-switching measures. Two sensitivity analyses repeated the rule-switching results with these sessions included and with every session below 65% overall accuracy excluded, the criterion of Wen et al. (2023; nine sessions). With the stricter criterion, 7 of 11 feedback-driven measures moved toward flexibility, as in the primary analysis; the stability composite's arm difference was Welch $t(19.7)=1.47$, $p=.16$, and its correlation with drinking change was $r_s=-.54$, $p=.009$.

Parametric Demand Selection Task. Responses not made by the deadline and presses of non-task keys were counted as errors and excluded from response-time measures. In the decision phase, the task re-presents a choice that was not made in time, so missed choices were dropped. In two post-dose sessions, decision-phase responses were inverted on 95–96% of answered trials, consistent with a reversed key mapping, and accuracy was scored with the mapping reversed. Baseline sessions whose learning-phase accuracy did not exceed chance (one-sided binomial test over 320 trials, $p \geq .05$; six participants) were excluded from all measures of the task, because deck–demand associations could not have been learned.

MSIT. The task file records the key pressed but not the mapping between keys and digits. The mapping was therefore inferred for each block as the rule (key equals the target digit, the target digit plus one, or the target position plus two) that reproduced at least 70% of the block's responses. Across all blocks, the best rule predicting different keys from the winning rule never matched more than 50% of responses, so the mapping was unambiguous; one block with a mid-session change of mapping could not be resolved and was not scored. In two post-dose sessions, 98–99% of interference trials were answered by the target's position rather than its identity. Because answering by position means the interference was not resolved, these sessions were excluded.

Operation span. Scores were computed from trial-level files as the partial-credit unit score. For one participant whose baseline trial-level file was not saved, the score was taken from the task's own summary log, which uses the same scoring; where both sources existed, they agreed to four decimal places. For one further participant, the baseline file contained no trials and no score was available.

Arm allocation. The dose table contained two rows with the same participant number and no row for another participant. The two rows were assigned using self-reported age and sex in the task files and baseline heavy-drinking days from the Timeline Followback. Both rows were in the psilocybin arm, so allocation itself was unaffected. The assignment was confirmed against the trial record, which also showed that one of these participants was taking an antidepressant (bupropion) that the dose table had not recorded; this was corrected.

#### Supplementary Results

Post-dose operation span. Fifteen participants completed operation span at both sessions (Table S8). Span did not change with practice (9.53 to 9.36; t(14)=−0.21, p=.84) and did not differ between arms at post-dose testing (d=−0.14 [−1.29, 1.02]; t(12)=−0.26, p=.80). Baseline and post-dose scores were correlated ($r_s$=+.63, p=.011).

Per-level demand selection models. Response time and accuracy in the learning phase changed with demand level (Table S9). The arms differed in how strongly RT and accuracy changed with demand level across both sessions (arm × level), but this difference did not change after drug administration (arm × level × session), and the arm × session interaction was not significant for any measure. Because the probabilities of choosing each deck sum across decks, the arm and session terms of the choice model are zero by construction; the arm × level × session term tests whether the drug changed how choice varied with demand (F(1, 144)=1.56, p=.21).

#### Table S1. Arm effects on all retest measures

ANCOVA of post-dose score on baseline score and arm. d = adjusted difference in pooled-SD units (psilocybin − placebo). Within-arm t from paired t-tests (n − 1 df); ANCOVA t has n − 3 df. q = BH-FDR across 34 measures. BF10 = Bayes factor for a drug effect (ANCOVA model with arm vs baseline-only model, default priors); BF01 = 1/BF10. Values between 1/3 and 3 are inconclusive. No measure reached BF01 > 3. Pole: S, higher = more stable; F, higher = more flexible (Table 1). Rule-switching measures exclude the three baseline sessions and one post-dose session whose accuracy did not exceed chance; with these sessions included, the transfer-phase switch cost was d=−0.75 [−1.72, 0.23], p=.12, transfer-phase post-switch accuracy d=+0.93 [−0.22, 2.07], p=.10, and 8 rather than 7 of the 11 feedback-driven measures moved toward flexibility. Demand selection measures exclude the six baseline sessions at chance in the learning phase. Script: code/10_drug_effects_and_coupling.R and code/14_bayes_factors.R (BF10); sensitivity analysis: code/15_sensitivity_rule_switching.R.

| Task | Measure | Pole | n P/S | Placebo pre → post | Psilocybin pre → post | Within-arm t (p), P / S | d [95% CI] | t (p) | q | BF10 |
|---|---|---|---|---|---|---|---|---|---|---|
| PRL | Reward-learning accuracy | – | 11/11 | 0.761 → 0.781 | 0.873 → 0.860 | +0.22 (.828) / −0.41 (.693) | +0.16 [−0.79, +1.12] | +0.36 (.723) | .993 | 0.43 |
| PRL | Punishment- | – | 11/1 | 0.732 → | 0.669 → | +4.24 | −0.00 [−0.72, | −0.0 | .993 | 0.40 |

| Task | Measure | Pole | n P/S | Placebo pre → post | Psilocybin pre → post | Within-arm t (p), P / S | d [95% CI] | t (p) | q | BF10 |
|---|---|---|---|---|---|---|---|---|---|---|
| | learning accuracy | | 1 | 0.804 | 0.747 | (.002) / +1.85 (.095) | +0.71] | 1 (.993) | | |
| PRL | Learning rate (gain) | F | 11/11 | 0.262 → 0.228 | 0.204 → 0.257 | −0.30 (.771) / +0.48 (.645) | +0.11 [−0.87, +1.08] | +0.23 (.821) | .993 | 0.39 |
| PRL | Learning rate (loss) | F | 11/11 | 0.496 → 0.374 | 0.423 → 0.478 | −0.75 (.472) / +0.36 (.730) | +0.28 [−0.61, +1.17] | +0.66 (.519) | .993 | 0.46 |
| PRL | Inverse temperature (gain) | S | 11/11 | 7.593 → 6.935 | 8.741 → 7.768 | −0.55 (.596) / −0.89 (.392) | +0.12 [−0.86, +1.09] | +0.25 (.806) | .993 | 0.41 |
| PRL | Inverse temperature (loss) | S | 11/11 | 4.745 → 5.628 | 4.578 → 5.039 | +0.59 (.571) / +0.26 (.802) | −0.15 [−1.08, +0.78] | −0.33 (.743) | .993 | 0.40 |
| PRL | Perseveration | S | 11/11 | 0.225 → 0.988 | 0.142 → 0.449 | +3.04 (.012) / +1.79 (.104) | −0.79 [−1.77, +0.19] | −1.69 (.107) | .703 | 1.12 |
| Rule switching | Overall accuracy | – | 6/7 | 0.770 → 0.758 | 0.779 → 0.805 | −0.70 (.513) / +1.41 (.208) | +0.68 [−0.33, +1.69] | +1.51 (.162) | .703 | 0.91 |
| Rule switching | Switch cost (learning) | S | 6/7 | 0.411 → 0.422 | 0.438 → 0.629 | +0.05 (.959) / +1.35 (.227) | +0.74 [−0.42, +1.89] | +1.42 (.186) | .703 | 0.89 |
| Rule switching | Switch cost (transfer) | S | 6/7 | 0.347 → 0.480 | 0.566 → 0.520 | +2.99 (.031) / −0.66 (.535) | −0.48 [−1.61, +0.65] | −0.95 (.365) | .867 | 0.55 |

| Task | Measure | Pole | n P/S | Placebo pre → post | Psilocybin pre → post | Within-arm t (p), P / S | d [95% CI] | t (p) | q | BF10 |
|---|---|---|---|---|---|---|---|---|---|---|
| Rule switching | Pre-switch accuracy (learning) | S | 6/7 | 0.811 → 0.756 | 0.838 → 0.933 | −0.39 (.714) / +1.16 (.290) | +0.98 [−0.25, +2.20] | +1.77 (.107) | .703 | 1.23 |
| Rule switching | Post-switch accuracy (learning) | F | 6/7 | 0.400 → 0.333 | 0.400 → 0.305 | −0.61 (.567) / −1.37 (.220) | −0.18 [−1.47, +1.11] | −0.31 (.763) | .993 | 0.47 |
| Rule switching | Pre-switch accuracy (transfer) | S | 6/7 | 0.833 → 0.867 | 0.971 → 0.937 | +0.85 (.434) / −1.16 (.289) | −0.25 [−1.47, +0.97] | −0.46 (.654) | .993 | 0.52 |
| Rule switching | Post-switch accuracy (transfer) | F | 6/7 | 0.487 → 0.387 | 0.406 → 0.417 | −2.13 (.087) / +0.26 (.805) | +0.79 [−0.43, +2.01] | +1.45 (.178) | .703 | 0.72 |
| Rule switching | Median RT (learning), ms | – | 6/7 | 960 → 1036 | 1084 → 1036 | +0.74 (.494) / −0.42 (.686) | −0.06 [−1.35, +1.24] | −0.10 (.924) | .993 | 0.45 |
| Rule switching | Median RT (transfer), ms | – | 6/7 | 1160 → 1215 | 1206 → 1126 | +0.54 (.610) / −0.74 (.488) | −0.47 [−1.56, +0.61] | −0.98 (.352) | .867 | 0.64 |
| Demand selection | Learning-phase accuracy | – | 7/9 | 0.807 → 0.885 | 0.855 → 0.833 | +1.33 (.232) / −0.58 (.576) | −0.58 [−1.61, +0.45] | −1.22 (.243) | .825 | 0.67 |
| Demand selection | Learning-phase RT, s | – | 7/9 | 0.637 → 0.591 | 0.733 → 0.775 | −1.15 (.293) / +1.52 (.167) | +1.01 [+0.30, +1.73] | +3.09 (.009) | .295 | 7.67 |
| Demand selecti | Task-switch cost (accuracy; | S | 7/9 | 0.063 → 0.041 | 0.068 → 0.091 | −0.66 (.532) / +0.77 | +0.80 [−0.32, +1.93] | +1.54 (.147 | .703 | 0.97 |

| Task | Measure | Pole | n P/S | Placebo pre → post | Psilocybin pre → post | Within-arm t (p), P / S | d [95% CI] | t (p) | q | BF10 |
|---|---|---|---|---|---|---|---|---|---|---|
| on | learning) | | | | | (.462) | | ) | | |
| Demand selection | Task-switch cost (RT; learning), s | S | 7/9 | 0.173 → 0.148 | 0.199 → 0.253 | −0.79 (.458) / +1.70 (.128) | +0.89 [+0.07, +1.71] | +2.35 (.035) | .466 | 2.36 |
| Demand selection | Decision-phase accuracy | – | 6/9 | 0.858 → 0.911 | 0.933 → 0.922 | +0.56 (.599) / −1.43 (.190) | +0.01 [−1.02, +1.04] | +0.02 (.985) | .993 | 0.45 |
| Demand selection | Decision-phase RT, s | – | 6/9 | 0.542 → 0.532 | 0.653 → 0.658 | −0.45 (.675) / +0.18 (.864) | +0.30 [−0.51, +1.10] | +0.80 (.438) | .930 | 0.67 |
| Demand selection | Task-switch cost (accuracy; decision) | S | 6/9 | 0.030 → 0.020 | 0.041 → 0.047 | −0.32 (.761) / +0.46 (.656) | +0.62 [−0.69, +1.93] | +1.03 (.324) | .867 | 0.66 |
| Demand selection | Task-switch cost (RT; decision), s | S | 6/9 | 0.127 → 0.125 | 0.173 → 0.173 | −0.12 (.911) / +0.01 (.992) | +0.17 [−0.67, +1.01] | +0.44 (.666) | .993 | 0.49 |
| Demand selection | P(choose least demanding) | – | 6/9 | 0.454 → 0.444 | 0.617 → 0.556 | −0.08 (.942) / −0.50 (.632) | +1.31 [+0.06, +2.55] | +2.29 (.041) | .466 | 1.31 |
| Demand selection | P(choose most demanding) | – | 6/9 | 0.546 → 0.444 | 0.410 → 0.488 | −1.07 (.333) / +0.58 (.580) | −0.05 [−1.42, +1.32] | −0.08 (.937) | .993 | 0.45 |
| Demand selection | Mean chosen demand level | – | 6/9 | 2.616 → 2.440 | 2.316 → 2.444 | −0.88 (.420) / +0.77 (.464) | −0.37 [−1.80, +1.07] | −0.56 (.588) | .993 | 0.49 |
| Demand | Selection RT, s | – | 6/9 | 0.860 → 0.754 | 0.859 → 0.818 | −1.81 (.130) / | +0.30 [−0.38, +0.98] | +0.97 | .867 | 0.59 |

| Task | Measure | Pole | n P/S | Placebo pre → post | Psilocybin pre → post | Within-arm t (p), P / S | d [95% CI] | t (p) | q | BF10 |
|---|---|---|---|---|---|---|---|---|---|---|
| selection | | | | | | −0.90 (.392) | | (.352) | | |
| MSIT | Control accuracy | – | 9/10 | 0.990 → 0.995 | 0.944 → 0.996 | +1.17 (.276) / +1.47 (.176) | +0.01 [−0.11, +0.12] | +0.15 (.883) | .993 | 0.42 |
| MSIT | Interference accuracy | – | 9/10 | 0.929 → 0.957 | 0.907 → 0.960 | +1.94 (.088) / +1.44 (.185) | +0.05 [−0.48, +0.58] | +0.21 (.839) | .993 | 0.41 |
| MSIT | Control RT, ms | – | 9/10 | 611 → 579 | 611 → 568 | −1.39 (.201) / −3.43 (.007) | −0.11 [−0.57, +0.34] | −0.53 (.604) | .993 | 0.45 |
| MSIT | Interference RT, ms | – | 9/10 | 914 → 850 | 935 → 862 | −2.27 (.053) / −3.98 (.003) | −0.03 [−0.61, +0.56] | −0.10 (.924) | .993 | 0.41 |
| MSIT | Interference effect (RT), ms | F | 9/10 | 302 → 271 | 325 → 294 | −1.40 (.198) / −1.41 (.193) | +0.20 [−0.72, +1.12] | +0.46 (.651) | .993 | 0.45 |
| MSIT | Interference effect (accuracy) | F | 9/10 | 0.060 → 0.038 | 0.036 → 0.036 | −1.90 (.094) / −0.04 (.967) | +0.35 [−0.47, +1.17] | +0.90 (.382) | .867 | 0.50 |

#### Table S2. Cognitive change and clinical change, all measures

Spearman $r_s$ between change in each measure (post-dose − baseline) and change in the proportion of drinking days (30 days after drug administration − baseline), pooled across arms. Arm-adjusted t from rank-based regression (n − 3 df); q = BH-FDR across 34. Heavy-day $r_s$ uses heavy-drinking days. PHQ β and t from rank regression adjusted for arm (n − 3 df). Rule-switching measures exclude sessions at chance. Script: code/10_drug_effects_and_coupling.R.

| Task | Measure | Pole | n | Drinking days $r_s$ (p) | Arm-adjusted t (p) | q | Heavy days $r_s$ (p) | PHQ β, t (p) |
|---|---|---|---|---|---|---|---|---|

| Task | Measure | Pole | n | Drinking days $r_s$ (p) | Arm-adjusted t (p) | q | Heavy days $r_s$ (p) | PHQ β, t (p) |
|---|---|---|---|---|---|---|---|---|
| PRL | Reward-learning accuracy | – | 22 | −0.48 (.022) | −2.27 (.035) | .494 | −0.37 (.090) | −0.25, −1.48 (.157) |
| PRL | Punishment-learning accuracy | – | 22 | −0.06 (.785) | −0.40 (.695) | .940 | +0.01 (.970) | −0.07, −0.41 (.683) |
| PRL | Learning rate (gain) | F | 22 | +0.36 (.095) | +1.75 (.097) | .494 | +0.24 (.283) | +0.23, +1.34 (.197) |
| PRL | Learning rate (loss) | F | 22 | +0.17 (.442) | +0.52 (.609) | .735 | +0.36 (.100) | +0.18, +1.06 (.302) |
| PRL | Inverse temperature (gain) | S | 22 | −0.37 (.087) | −1.81 (.087) | .494 | −0.21 (.354) | −0.26, −1.56 (.136) |
| PRL | Inverse temperature (loss) | S | 22 | −0.17 (.448) | −0.70 (.494) | .735 | −0.30 (.175) | −0.22, −1.30 (.209) |
| PRL | Perseveration | S | 22 | −0.46 (.031) | −2.11 (.048) | .494 | −0.30 (.168) | −0.11, −0.64 (.530) |
| Rule switching | Overall accuracy | – | 13 | −0.07 (.809) | −1.39 (.194) | .940 | +0.19 (.541) | +0.00, +0.01 (.992) |
| Rule switching | Switch cost (learning) | S | 13 | −0.36 (.233) | −2.16 (.056) | .565 | −0.34 (.257) | +0.26, +1.12 (.289) |
| Rule switching | Switch cost (transfer) | S | 13 | −0.47 (.106) | −0.64 (.534) | .494 | −0.44 (.136) | −0.39, −1.39 (.195) |

| Task | Measure | Pole | n | Drinking days $r_s$ (p) | Arm-adjusted t (p) | q | Heavy days $r_s$ (p) | PHQ β, t (p) |
|---|---|---|---|---|---|---|---|---|
| Rule switching | Pre-switch accuracy (learning) | S | 13 | −0.45 (.127) | −2.89 (.016) | .494 | −0.37 (.212) | +0.36, +1.61 (.139) |
| Rule switching | Post-switch accuracy (learning) | F | 13 | +0.13 (.678) | +0.58 (.572) | .933 | +0.22 (.473) | −0.08, −0.31 (.765) |
| Rule switching | Pre-switch accuracy (transfer) | S | 13 | −0.38 (.203) | −0.75 (.473) | .530 | −0.03 (.927) | −0.36, −1.49 (.166) |
| Rule switching | Post-switch accuracy (transfer) | F | 13 | +0.39 (.183) | +0.65 (.528) | .517 | +0.55 (.053) | +0.22, +0.83 (.425) |
| Rule switching | Median RT (learning) | – | 13 | +0.29 (.334) | +1.41 (.188) | .709 | +0.22 (.470) | −0.34, −1.52 (.159) |
| Rule switching | Median RT (transfer) | – | 13 | +0.41 (.167) | +2.91 (.015) | .517 | +0.52 (.074) | −0.06, −0.25 (.804) |
| Demand selection | Learning-phase accuracy | – | 16 | −0.39 (.131) | −1.27 (.226) | .494 | −0.32 (.221) | −0.15, −0.65 (.528) |
| Demand selection | Learning-phase RT | – | 16 | +0.35 (.181) | +1.01 (.329) | .517 | +0.35 (.184) | +0.07, +0.28 (.788) |
| Demand | Task-switch | S | 16 | −0.10 | −0.66 | .940 | −0.15 | +0.10, +0.43 |

| Task | Measure | Pole | n | Drinking days $r_s$ (p) | Arm-adjusted t (p) | q | Heavy days $r_s$ (p) | PHQ β, t (p) |
|---|---|---|---|---|---|---|---|---|
| selection | cost (accuracy; learning) | | | (.720) | (.518) | | (.575) | (.671) |
| Demand selection | Task-switch cost (RT; learning) | S | 16 | +0.18 (.502) | +0.24 (.812) | .749 | +0.06 (.822) | −0.12, −0.48 (.636) |
| Demand selection | Decision-phase accuracy | – | 15 | −0.28 (.314) | −0.79 (.445) | .709 | −0.21 (.442) | −0.09, −0.52 (.612) |
| Demand selection | Decision-phase RT | – | 15 | +0.43 (.112) | +1.50 (.160) | .494 | +0.46 (.086) | −0.04, −0.24 (.811) |
| Demand selection | Task-switch cost (accuracy; decision) | S | 15 | −0.24 (.393) | −1.15 (.272) | .735 | −0.11 (.686) | −0.17, −1.07 (.307) |
| Demand selection | Task-switch cost (RT; decision) | S | 15 | +0.21 (.454) | +0.69 (.504) | .735 | +0.21 (.450) | −0.37, −2.98 (.011) |
| Demand selection | P(choose least demanding) | – | 15 | +0.06 (.830) | +0.17 (.868) | .940 | +0.10 (.714) | +0.14, +0.87 (.400) |
| Demand selection | P(choose most demanding) | – | 15 | +0.02 (.952) | −0.13 (.901) | .952 | −0.07 (.800) | −0.11, −0.67 (.514) |
| Demand selection | Mean chosen demand | – | 15 | −0.09 (.746) | −0.41 (.690) | .940 | −0.22 (.439) | −0.19, −1.21 (.248) |

| Task | Measure | Pole | n | Drinking days $r_s$ (p) | Arm-adjusted t (p) | q | Heavy days $r_s$ (p) | PHQ β, t (p) |
|---|---|---|---|---|---|---|---|---|
| | level | | | | | | | |
| Demand selection | Selection RT | – | 15 | −0.04 (.899) | −0.27 (.792) | .952 | −0.03 (.913) | −0.15, −0.94 (.364) |
| MSIT | Control accuracy | – | 19 | +0.21 (.390) | +0.69 (.501) | .735 | +0.21 (.388) | +0.11, +0.62 (.542) |
| MSIT | Interference accuracy | – | 19 | +0.02 (.920) | +0.15 (.881) | .952 | −0.09 (.720) | +0.01, +0.04 (.969) |
| MSIT | Control RT | – | 19 | −0.47 (.044) | −1.98 (.065) | .494 | −0.46 (.050) | −0.15, −0.87 (.396) |
| MSIT | Interference RT | – | 19 | −0.10 (.686) | −0.37 (.713) | .933 | −0.12 (.616) | +0.05, +0.28 (.785) |
| MSIT | Interference effect (RT) | F | 19 | +0.16 (.507) | +0.64 (.531) | .749 | +0.10 (.678) | +0.20, +1.25 (.230) |
| MSIT | Interference effect (accuracy) | F | 19 | +0.02 (.934) | −0.10 (.919) | .952 | +0.07 (.788) | +0.15, +0.90 (.380) |

#### Table S3. Baseline cognition and baseline drinking, all measures

Spearman $r_s$ (p) between baseline cognition and baseline drinking rates (90-day window). None survives BH-FDR across 105 tests (smallest q=.41).

| Task | Measure | n | Drinking days | Heavy days | Drinks per drinking day |
|---|---|---|---|---|---|
| PRL | Reward-learning accuracy | 24 | −0.42 (.041) | −0.21 (.332) | +0.27 (.201) |

| Task | Measure | n | Drinking days | Heavy days | Drinks per drinking day |
|---|---|---|---|---|---|
| PRL | Punishment-learning accuracy | 24 | −0.02 (.932) | −0.25 (.245) | +0.14 (.526) |
| PRL | Learning rate (gain) | 24 | −0.09 (.664) | −0.44 (.031) | −0.36 (.084) |
| PRL | Learning rate (loss) | 24 | −0.13 (.554) | +0.16 (.466) | +0.23 (.271) |
| PRL | Inverse temperature (gain) | 24 | −0.24 (.252) | −0.02 (.917) | +0.18 (.402) |
| PRL | Inverse temperature (loss) | 24 | +0.06 (.795) | −0.28 (.182) | −0.14 (.506) |
| PRL | Perseveration | 24 | −0.42 (.043) | +0.17 (.421) | +0.57 (.004) |
| Rule switching | Overall accuracy | 17 | −0.06 (.833) | −0.22 (.386) | +0.12 (.652) |
| Rule switching | Switch cost (learning) | 17 | −0.18 (.491) | +0.00 (.996) | +0.41 (.100) |
| Rule switching | Switch cost (transfer) | 17 | −0.11 (.683) | +0.07 (.785) | +0.52 (.032) |
| Rule switching | Pre-switch accuracy (learning) | 17 | −0.22 (.405) | −0.34 (.188) | +0.18 (.487) |
| Rule switching | Post-switch accuracy (learning) | 17 | +0.08 (.773) | −0.29 (.263) | −0.49 (.044) |
| Rule switching | Pre-switch accuracy (transfer) | 17 | −0.44 (.078) | +0.07 (.786) | +0.49 (.045) |
| Rule switching | Post-switch accuracy (transfer) | 17 | −0.09 (.726) | −0.09 (.726) | −0.35 (.167) |
| Rule switching | Median RT | 17 | −0.11 (.673) | +0.06 (.808) | −0.10 (.708) |

| Task | Measure | n | Drinking days | Heavy days | Drinks per drinking day |
|---|---|---|---|---|---|
| | (learning) | | | | |
| Rule switching | Median RT (transfer) | 17 | −0.06 (.831) | +0.08 (.772) | −0.06 (.831) |
| Demand selection | Learning-phase accuracy | 19 | +0.24 (.319) | −0.31 (.199) | −0.11 (.652) |
| Demand selection | Learning-phase RT | 19 | −0.02 (.920) | −0.11 (.668) | −0.32 (.182) |
| Demand selection | Task-switch cost (accuracy; learning) | 19 | −0.21 (.398) | +0.12 (.613) | +0.13 (.591) |
| Demand selection | Task-switch cost (RT; learning) | 19 | +0.30 (.213) | +0.25 (.305) | −0.16 (.508) |
| Demand selection | Decision-phase accuracy | 18 | +0.13 (.618) | −0.10 (.685) | +0.20 (.428) |
| Demand selection | Decision-phase RT | 18 | −0.09 (.729) | +0.06 (.807) | −0.06 (.818) |
| Demand selection | Task-switch cost (accuracy; decision) | 18 | −0.29 (.238) | −0.09 (.723) | +0.15 (.547) |
| Demand selection | Task-switch cost (RT; decision) | 18 | +0.38 (.123) | +0.24 (.332) | −0.29 (.239) |
| Demand selection | P(choose least demanding) | 18 | −0.48 (.044) | −0.34 (.165) | −0.08 (.766) |
| Demand selection | P(choose most demanding) | 18 | +0.30 (.220) | +0.32 (.199) | −0.05 (.835) |
| Demand selection | Mean chosen demand level | 18 | +0.36 (.147) | +0.29 (.250) | +0.01 (.958) |
| Demand selection | Selection RT | 18 | −0.27 (.285) | −0.40 (.096) | −0.00 (.993) |

| Task | Measure | n | Drinking days | Heavy days | Drinks per drinking day |
|---|---|---|---|---|---|
| MSIT | Control accuracy | 23 | +0.24 (.280) | −0.02 (.922) | −0.39 (.069) |
| MSIT | Interference accuracy | 23 | +0.08 (.734) | +0.23 (.301) | +0.22 (.323) |
| MSIT | Control RT | 23 | −0.26 (.221) | +0.07 (.752) | +0.42 (.047) |
| MSIT | Interference RT | 23 | −0.27 (.211) | −0.25 (.249) | +0.18 (.410) |
| MSIT | Interference effect (RT) | 23 | +0.12 (.579) | −0.36 (.087) | −0.55 (.008) |
| MSIT | Interference effect (accuracy) | 23 | −0.09 (.667) | −0.45 (.031) | −0.40 (.061) |
| Operation span | Partial-credit span | 18 | −0.42 (.080) | −0.36 (.137) | +0.13 (.610) |

##### Table S4. Working-memory capacity and drinking change: robustness checks

Outcome: change in percent heavy-drinking days and percent drinking days from baseline to the first 30-day follow-up window, unless noted. Primary model: rank-based partial regression of drinking change on baseline operation span (OSpan), adjusting for arm and baseline drinking. Covariate models are linear (drinking change ~ OSpan + covariate), so their β is in raw OSpan units.

| Analysis | n | Heavy-drinking days | Drinking days |
|---|---|---|---|
| Primary (rank-based; arm + baseline adjusted) | 18 | β=+0.57, t(14)=3.94, p=.0015 | β=+0.63, t(14)=3.76, p=.0021 |
| FDR across 70 prognostic tests | 18 | q=.074 | q=.074 |
| 90-day window | 17 | t(13)=4.00, p=.0015 | t(13)=4.27, p=.0009 |
| Excluding the baseline-only participant | 17 | β=+0.57, t(13)=3.82, p=.0021 | β=+0.62, t(13)=3.69, p=.0027 |
| Leave-one-out (largest p across 18 samples) | 17 | smallest t=3.00, p=.010 | smallest t=3.20, p=.007 |
| Pearson (linear; arm + | 18 | t(14)=3.95, p=.0015 | t(14)=2.84, p=.013 |

| Analysis | n | Heavy-drinking days | Drinking days |
|---|---|---|---|
| baseline adjusted) | | | |
| Processing accuracy ≥85% only | 11 | β=+0.56, t(7)=1.99, p=.087 | β=+0.83, t(7)=2.97, p=.021 |
| Placebo arm only (Spearman) | 10 | $r_s$=.67, p=.033 | $r_s$=.70, p=.025 |
| Psilocybin arm only (Spearman) | 8 | $r_s$=.90, p=.002 | $r_s$=.83, p=.011 |
| Arm × OSpan interaction | 18 | t(13)=1.61, p=.13 | t(13)=1.03, p=.32 |
| + age (linear) | 18 | t(13)=4.26, p=.0009 | t(13)=2.90, p=.012 |
| + sex (linear) | 18 | t(13)=4.09, p=.0013 | t(13)=2.73, p=.017 |
| + screening QIDS-SR (linear) | 18 | t(13)=3.60, p=.0032 | t(13)=2.27, p=.041 |
| + antidepressant use (linear) | 18 | t(13)=3.89, p=.0019 | t(13)=2.45, p=.029 |

OSpan was unrelated to age (r=.05) and screening QIDS-SR (r=−.17). Script: code/12_prediction_and_wmc.R.

##### Table S5. Willingness to exert control (demand avoidance) and PRISM's predictions

Demand avoidance = proportion of free choices of the less demanding deck (72 choices; 0.5 = indifference). Baseline sessions at chance in the learning phase are excluded unless stated. Script: code/13_demand_avoidance.R.

| Test | Statistic | n |
|---|---|---|
| Baseline mean vs indifference | .54 vs .50, t(17)=1.14, p=.27 | 18 |
| × operation span | $r_s$=+.10, p=.75 | 13 |
| × demand selection accuracy, RT, switch costs (performance, same task) | all $\lvert r_s \rvert$≤.36, all p≥.14; BF01 0.9–2.0 | 18 |
| × rule-switching cost | $r_s$=+.48, p=.12 | 12 |
| × PHQ, same visit | $r_s$=+.71, p=.001 | 17 |
| × PHQ, same visit, chance-level sessions included | $r_s$=+.68, p<.001 | 22 |
| × QIDS-SR at screening | $r_s$=+.06, p=.83 | 18 |

| Test | Statistic | n |
|---|---|---|
| Post-dose avoidance × post-dose PHQ (all post-dose sessions) | $r_s$=−.16, p=.51 | 20 |
| Test–retest correlation, within arm (chance-level sessions included: $r_s$=−.13, n=20) | $r_s$=−.27, p=.33 (without removing arm means: +.03, p=.91) | 15 |
| Arm effect at post-dose testing (ANCOVA) | d=+0.47 [−1.06, 2.00], t(12)=0.67, p=.52; placebo .43→.53, psilocybin .60→.53; r(baseline, change)=−.77 | 6/9 |
| Change × drinking-day change | $r_s$=−.02, p=.94 | 15 |
| Change × PHQ change | $r_s$=+.27, p=.33 | 15 |
| Baseline avoidance → drinking-day change (× arm) | adjusted β=−0.08, t(14)=−0.33, p=.74, BF01=2.3; interaction t(13)=0.54, p=.60, BF01=1.8 | 18 |
| Baseline avoidance → heavy-day change (× arm) | adjusted β=−0.32, t(14)=−1.48, p=.16, BF01=2.0; interaction t(13)=0.32, p=.75, BF01=2.6 | 18 |
| Baseline avoidance → PHQ change (× arm) | adjusted β=+0.08, t(13)=0.64, p=.54, BF01=3.4; interaction t(12)=−0.47, p=.64, BF01=3.3; unadjusted $r_s$=−.61 (explained by baseline PHQ); psilocybin arm unadjusted $r_s$=−.50, p=.14 | 17 |

#### Table S6. Baseline construct validation of the stability–flexibility classification

Baseline session only (before randomization). Each index is aligned so that higher values indicate greater stability; flexibility-pole indices (F) are sign-reversed. Rule-switching and demand selection indices exclude sessions at chance. Mean Spearman $r_s$ across the 23 cross-task pairs = +.11; permutation p =.032 (10,000 permutations, participants shuffled independently within each index); 16 of 23 pairs positive. Within-task pairs are shown for completeness and were not part of the test. Only 12 participants contributed all eight indices, so factor analysis was not attempted. Script: code/21_construct_validation.py.

*A. Correlations between aligned indices*

| Index 1 | Index 2 | Pair type | n | $r_s$ | p |
|---|---|---|---|---|---|
| Cued switch cost, RT (S) | Cued switch cost, accuracy (S) | Within-task | 19 | +.24 | .318 |

| Index 1 | Index 2 | Pair type | n | $r_s$ | p |
|---|---|---|---|---|---|
| PRL perseveration (S) | PRL learning rate (F, reversed) | Within-task | 24 | +.22 | .308 |
| PRL perseveration (S) | PRL inverse temperature (S) | Within-task | 24 | +.35 | .093 |
| Rule switching: pre-switch accuracy (S) | Rule switching: post-switch accuracy (F, reversed) | Within-task | 17 | +.47 | .055 |
| PRL inverse temperature (S) | PRL learning rate (F, reversed) | Within-task | 24 | +.67 | <.001 |
| PRL learning rate (F, reversed) | Cued switch cost, accuracy (S) | Cross-task | 19 | -.00 | .989 |
| Rule switching: pre-switch accuracy (S) | Cued switch cost, accuracy (S) | Cross-task | 12 | -.11 | .737 |
| PRL perseveration (S) | Rule switching: post-switch accuracy (F, reversed) | Cross-task | 16 | +.11 | .696 |
| PRL inverse temperature (S) | Cued switch cost, RT (S) | Cross-task | 19 | +.11 | .667 |
| Rule switching: post-switch accuracy (F, reversed) | Cued switch cost, RT (S) | Cross-task | 12 | -.16 | .609 |
| PRL learning rate (F, reversed) | Rule switching: pre-switch accuracy (S) | Cross-task | 16 | +.15 | .583 |
| PRL perseveration (S) | Rule switching: pre-switch accuracy (S) | Cross-task | 16 | +.16 | .542 |
| Rule switching: pre-switch accuracy (S) | MSIT interference, RT (F, reversed) | Cross-task | 17 | +.16 | .532 |
| PRL inverse temperature (S) | Rule switching: post-switch accuracy (F, reversed) | Cross-task | 16 | +.18 | .504 |
| PRL inverse temperature (S) | Cued switch cost, accuracy (S) | Cross-task | 19 | +.17 | .485 |
| PRL learning rate (F, reversed) | Cued switch cost, RT (S) | Cross-task | 19 | +.20 | .416 |
| Cued switch cost, RT (S) | MSIT interference, RT (F, reversed) | Cross-task | 17 | -.23 | .384 |
| PRL perseveration (S) | Cued switch cost, RT (S) | Cross-task | 19 | -.21 | .379 |
| PRL perseveration (S) | Cued switch cost, accuracy (S) | Cross-task | 19 | +.24 | .325 |

| Index 1 | Index 2 | Pair type | n | $r_s$ | p |
|---|---|---|---|---|---|
| Rule switching: post-switch accuracy (F, reversed) | Cued switch cost, accuracy (S) | Cross-task | 12 | -.31 | .324 |
| PRL learning rate (F, reversed) | Rule switching: post-switch accuracy (F, reversed) | Cross-task | 16 | +.27 | .321 |
| PRL inverse temperature (S) | Rule switching: pre-switch accuracy (S) | Cross-task | 16 | +.29 | .277 |
| PRL inverse temperature (S) | MSIT interference, RT (F, reversed) | Cross-task | 22 | +.27 | .233 |
| PRL learning rate (F, reversed) | MSIT interference, RT (F, reversed) | Cross-task | 22 | +.33 | .132 |
| Rule switching: pre-switch accuracy (S) | Cued switch cost, RT (S) | Cross-task | 12 | -.49 | .108 |
| Rule switching: post-switch accuracy (F, reversed) | MSIT interference, RT (F, reversed) | Cross-task | 17 | +.42 | .095 |
| Cued switch cost, accuracy (S) | MSIT interference, RT (F, reversed) | Cross-task | 17 | +.49 | .047 |
| PRL perseveration (S) | MSIT interference, RT (F, reversed) | Cross-task | 22 | +.57 | .006 |

*B. Test–retest reliability (Spearman $r_s$ between baseline and post-dose scores after removing arm means)*

| Index | n | $r_s$ | p |
|---|---|---|---|
| PRL perseveration (S) | 22 | +.12 | .584 |
| PRL inverse temperature (S) | 22 | +.25 | .266 |
| PRL learning rate (F, reversed) | 22 | -.06 | .793 |
| Rule switching: pre-switch accuracy (S) | 13 | +.42 | .157 |
| Rule switching: post-switch accuracy (F, reversed) | 13 | -.01 | .986 |
| Cued switch cost, RT (S) | 16 | +.45 | .083 |
| Cued switch cost, accuracy (S) | 16 | +.18 | .506 |
| MSIT interference, RT (F, reversed) | 19 | +.40 | .091 |

#### Table S7. Effect sizes and Bayes factors for the main claims

Effect sizes with 95% confidence intervals and Bayes factors (BayesFactor 0.9.12-4.8, default priors). Drug effects: ANCOVA model including arm compared with the baseline-only model (d, baseline-adjusted difference in pooled-SD units; composites: Cohen's d from Welch comparison of change). Correlations: Bayes factors on ranks with the default stretched-beta prior; confidence intervals use the Bonett–Wright approximation. Rank regressions: models with and without the predictor, ranks of all continuous variables. Evidence categories follow Lee and Wagenmakers (2013): BF between 1/3 and 3, inconclusive; 3–10, moderate; above 10, strong. Pilot effect sizes are imprecise and should not be used directly for power calculations (Leon et al., 2011). Script: code/14_bayes_factors.R; composite and clinical effect sizes from code/10_drug_effects_and_coupling.R and code/11_sample_and_clinical.R.

| Claim | Effect size [95% CI] | n | BF10 | BF01 | Evidence |
|---|---|---|---|---|---|
| **Drug effects at post-dose testing (ANCOVA; d, psilocybin − placebo)** | | | | | |
| MSIT interference effect, RT | +0.20 [−0.72, 1.12] | 9/10 | 0.45 | 2.22 | Inconclusive |
| MSIT interference effect, accuracy | +0.35 [−0.47, 1.17] | 9/10 | 0.50 | 2.01 | Inconclusive |
| Reward-learning accuracy | +0.16 [−0.79, 1.12] | 11/11 | 0.43 | 2.30 | Inconclusive |
| Punishment-learning accuracy | −0.00 [−0.72, 0.71] | 11/11 | 0.40 | 2.52 | Inconclusive |
| Perseveration | −0.79 [−1.77, 0.19] | 11/11 | 1.12 | 0.89 | Inconclusive |
| Rule-switching cost, transfer | −0.48 [−1.61, 0.65] | 6/7 | 0.55 | 1.82 | Inconclusive |
| Post-switch accuracy, transfer | +0.79 [−0.43, 2.01] | 6/7 | 0.72 | 1.40 | Inconclusive |
| Learning-phase RT, demand selection | +1.01 [0.30, 1.73] | 7/9 | 7.67 | 0.13 | Moderate for effect |
| Switch-trial RT, learning phase | +1.12 [0.46, 1.77] | 7/9 | 17 | 0.06 | Strong for effect |
| Repeat-trial RT, learning phase | +0.64 [−0.08, 1.36] | 7/9 | 1.54 | 0.65 | Inconclusive |
| Task-switch cost in RT, learning phase | +0.89 [0.07, 1.71] | 7/9 | 2.36 | 0.42 | Inconclusive |
| Learning-phase accuracy, demand selection | −0.58 [−1.61, 0.45] | 7/9 | 0.67 | 1.49 | Inconclusive |
| P(choose least demanding deck) | +1.31 [0.06, 2.55] | 6/9 | 1.31 | 0.76 | Inconclusive |
| Demand avoidance | +0.47 [−1.06, 2.00] | 6/9 | 0.51 | 1.96 | Inconclusive |
| Stability composite (narrow) | −0.73 [−1.59, 0.14] | 11/11 | 1.05 | 0.95 | Inconclusive |
| Stability composite (broad) | −0.21 [−1.04, 0.63] | 11/11 | 0.42 | 2.39 | Inconclusive |
| PHQ | −0.88 [−1.62, −0.14] | 11/12 | 2.73 | 0.37 | Inconclusive |

| Claim | Effect size [95% CI] | n | BF10 | BF01 | Evidence |
|---|---|---|---|---|---|
| Proportion of drinking days, 30 days | +0.13 [−0.65, 0.92] | 13/12 | 0.39 | 2.57 | Inconclusive |
| Proportion of heavy-drinking days, 30 days | +0.15 [−0.39, 0.69] | 13/12 | 0.42 | 2.39 | Inconclusive |
| **Baseline associations (Spearman $r_s$, approximate 95% CI)** | | | | | |
| Demand avoidance vs indifference (.5) | mean .54 (SD .14) | 18 | 0.43 | 2.33 | Inconclusive |
| Demand avoidance × operation span | +.10 [−.48, +.62] | 13 | 0.58 | 1.72 | Inconclusive |
| Demand avoidance × PHQ, same visit | +.71 [+.30, +.90] | 17 | 24 | 0.04 | Strong for effect |
| Post-dose demand avoidance × post-dose PHQ | −.16 [−.56, +.31] | 20 | 0.56 | 1.78 | Inconclusive |
| Perseveration × rule-switching cost (transfer) | +.21 [−.33, +.64] | 16 | 0.65 | 1.54 | Inconclusive |
| Demand avoidance × 8 performance measures of the same task | $r_s$ from −.07 to +.36 | 18 | 0.50–1.13 | 0.88–2.01 | Inconclusive |
| **Cognitive change and clinical change (Spearman $r_s$, approximate 95% CI)** | | | | | |
| Stability composite change × drinking-day change | −.64 [−.85, −.27] | 22 | 29 | 0.03 | Strong for effect |
| Broad composite change × drinking-day change | −.35 [−.68, +.10] | 22 | 1.25 | 0.80 | Inconclusive |
| Perseveration change × drinking-day change | −.46 [−.75, −.02] | 22 | 2.94 | 0.34 | Inconclusive |
| Reward-learning accuracy change × drinking-day change | −.48 [−.76, −.05] | 22 | 3.68 | 0.27 | Moderate for effect |
| Stability composite change × PHQ change | +.04 [−.39, +.46] | 22 | 0.46 | 2.16 | Inconclusive |
| Demand avoidance change × drinking-day change | −.02 [−.53, +.50] | 15 | 0.53 | 1.88 | Inconclusive |
| Demand avoidance change × PHQ change | +.27 [−.29, +.69] | 15 | 0.76 | 1.32 | Inconclusive |
| PHQ change × drinking-day change | −.15 [−.53, +.28] | 23 | 0.55 | 1.83 | Inconclusive |

| Claim | Effect size [95% CI] | n | BF10 | BF01 | Evidence |
|---|---|---|---|---|---|
| Stability composite change × drinking-day change, arm-adjusted | rank regression | 22 | 16 | 0.06 | Strong for effect |
| Stability composite change × drinking-day change, participants with both measures | −.56 [−.86, +.04] | 13 | 2.21 | 0.45 | Inconclusive |
| **Prediction from baseline (rank-partial β, adjusted for arm and baseline outcome)** | | | | | |
| Operation span → heavy-drinking-day change | β=+0.57 | 18 | 25 | 0.04 | Strong for effect |
| Operation span → drinking-day change | β=+0.63 | 18 | 19 | 0.05 | Strong for effect |
| Operation span × arm (heavy-drinking days) | interaction | 18 | 0.76 | 1.32 | Inconclusive |
| Operation span × arm (drinking days) | interaction | 18 | 0.51 | 1.95 | Inconclusive |
| Operation span → PHQ change | β=−0.04 | 17 | 0.35 | 2.85 | Inconclusive |
| Demand avoidance → drinking-day change | β=−0.08 | 18 | 0.44 | 2.30 | Inconclusive |
| Demand avoidance → heavy-drinking-day change | β=−0.32 | 18 | 0.51 | 1.96 | Inconclusive |
| Demand avoidance → PHQ change | β=+0.08 | 17 | 0.30 | 3.39 | Moderate against |
| Demand avoidance × arm → PHQ change | interaction | 17 | 0.31 | 3.25 | Moderate against |

#### Table S8. Operation span at baseline and post-dose testing

Participants with operation span at both sessions. Drug effect: ANCOVA of the post-dose score on the baseline score and arm, d in pooled-SD units (psilocybin − placebo). Script: code/17_supplementary_analyses.R.

| Statistic | Value | n |
|---|---|---|
| Baseline → post-dose, placebo | 9.45 → 9.49 | 8 |
| Baseline → post-dose, psilocybin | 9.61 → 9.21 | 7 |
| Practice effect, both arms (paired t) | 9.53 → 9.36; t(14)=−0.21, p=.835 | 15 |

| Statistic | Value | n |
|---|---|---|
| Drug effect at post-dose testing (ANCOVA) | d=−0.14 [−1.29, 1.02]; t(12)=−0.26, p=.797 | 8/7 |
| Baseline–post-dose correlation (Spearman) | $r_s$=+.63, p=.011 | 15 |

##### Table S9. Per-level demand selection models

Linear mixed models of each measure at the four demand levels: value ~ arm × demand level × session + (1 + session | participant), with demand level as the centred task-switch probability (.13, .38, .63, .88). Satterthwaite F tests. Baseline sessions at chance in the learning phase are excluded. Choice probabilities sum across decks, so the arm and session terms of the choice model are zero by construction (singular fit). Script: code/17_supplementary_analyses.R.

| Term | Learning-phase RT (s) (n=24) | Learning-phase accuracy (n=24) | P(choose deck when offered) (n=23) |
|---|---|---|---|
| Arm | F(1, 20.8)=2.05, p=.167 | F(1, 17.4)=0.04, p=.838 | F(1, 144.0)=0.00, p>.999 |
| Demand level | F(1, 116.0)=318.12, p<.001 | F(1, 116.0)=35.72, p<.001 | F(1, 144.0)=1.61, p=.207 |
| Session | F(1, 16.0)=0.01, p=.929 | F(1, 13.7)=0.00, p=.977 | F(1, 144.0)=0.00, p>.999 |
| Arm × level | F(1, 116.0)=9.06, p=.003 | F(1, 116.0)=5.61, p=.020 | F(1, 144.0)=2.80, p=.096 |
| Arm × session | F(1, 16.0)=3.38, p=.084 | F(1, 13.7)=1.50, p=.241 | F(1, 144.0)=0.00, p>.999 |
| Level × session | F(1, 116.0)=0.51, p=.475 | F(1, 116.0)=0.13, p=.721 | F(1, 144.0)=0.19, p=.663 |
| Arm × level × session | F(1, 116.0)=0.86, p=.355 | F(1, 116.0)=0.18, p=.674 | F(1, 144.0)=1.56, p=.214 |

### Figure S1. Baseline construct validation

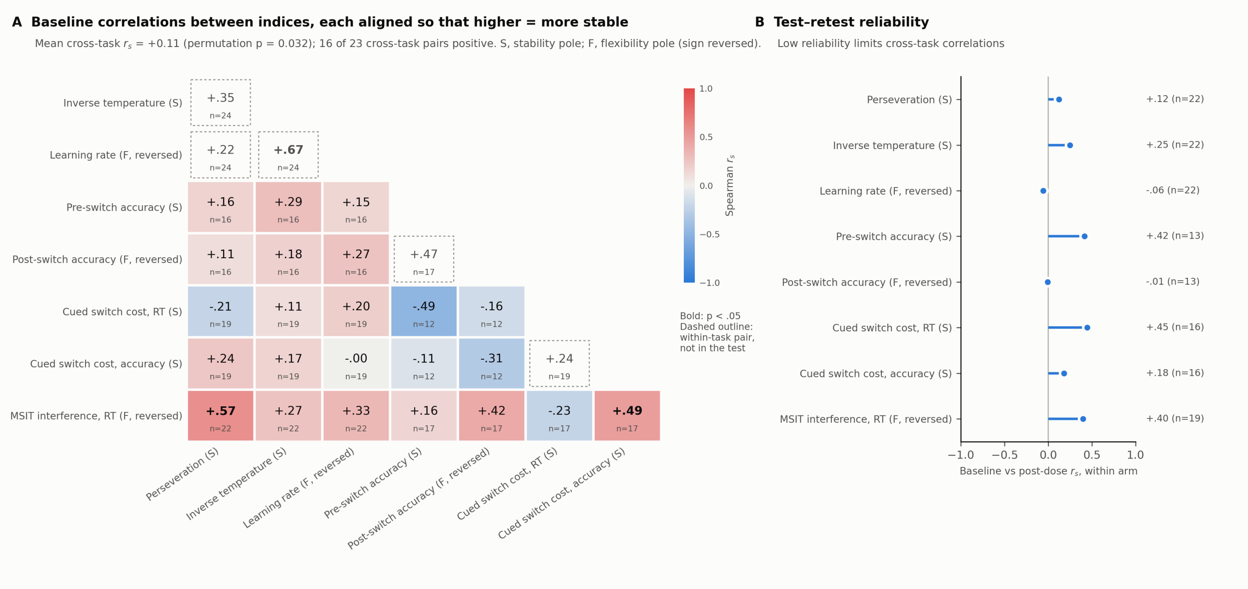


*Figure S1. Baseline construct validation of the stability–flexibility classification. (A) Spearman correlations between baseline indices, each aligned so that higher values indicate greater stability (S, stability pole; F, flexibility pole, sign-reversed). Coloured cells are cross-task pairs; dashed, unfilled cells are within-task pairs, which were not part of the test. Bold values, p<.05; n is given in each cell. (B) Test–retest reliability of each index (Spearman $r_s$ between baseline and post-dose scores after removing arm means). Low reliability limits the size of correlations between tasks.*
